# Calculating variant penetrance using family history of disease and population data Authorship

**DOI:** 10.1101/2021.03.16.21253691

**Authors:** Thomas P Spargo, Sarah Opie-Martin, Cathryn M Lewis, Alfredo Iacoangeli, Ammar Al-Chalabi

## Abstract

**Purpose:** Genetic penetrance is the probability of a phenotype manifesting if one harbours a specific pathogenic variant. For most Mendelian genes, penetrance is incomplete and may be age-dependent. Accurate penetrance estimates are important in many biomedical fields including genetic counselling, disease research, and for gene therapy. The main methods for its estimation are limited in situations where large family pedigrees are not available, the disease is rare, late onset, or complex.

**Methods:** Here we present a novel method for penetrance estimation in autosomal dominant phenotypes. It uses population-scale data regarding the distribution of a variant among unrelated people affected (cases) and unaffected (controls) by an associated phenotype and can be operated using samples of affected people only by considering family disease history.

**Results:** The method is validated within simulation studies. Candidate variant-disease case studies yield estimates which align with existing disease knowledge and estimates derived using established methods.

**Conclusion:** We have presented a valid method for penetrance estimation in autosomal dominant traits which avoids kinship-specific penetrance estimates and the ascertainment biases that can arise when sampling rare variants among control populations. It can be accessed from our public web server (https://adpenetrance.rosalind.kcl.ac.uk) and is available as an open-source R library (https://github.com/ThomasPSpargo/adpenetrance).

## Introduction

Penetrance is the probability of developing a specific trait given a genetic variant or set of variants. Some pathogenic variants are fully penetrant, and people harbouring them always develop the associated phenotype. For instance, a trinucleotide CAG repeat expansion within the *HTT* gene is fully penetrant for Huntington’s Disease by 80 years of age among people harbouring an expansion variant larger than 41 repeats^1^. For many variants however, penetrance is incomplete, and those with risk variants can remain unaffected throughout their life. For example, the p.Gly2019Ser variant of the *LRRK2* gene exhibits incomplete penetrance for Parkinson’s Disease (PD), meaning that it elevates risk but does not necessarily result in its manifestation^2^.

In medical genetics, estimating the penetrance of pathogenic variants is vital for the correct interpretation of genetic test results. This importance will increase as genome sequencing becomes routine, both within and outside clinical practice, alongside advancements in precision medicine and gene therapy^3-6^.

Several methods exist for penetrance estimation. The first and most widely used is based on statistical examination of how the variant segregates with the phenotype within pedigrees^7^. However, the generalisability of estimates derived from specific families may be limited. Other approaches examine the incidence of disease in a sample of unrelated people who harbour a variant^8, 9^. Without systematic sampling, these estimates can be affected by ascertainment bias. Where large pedigrees are not available, or if disease is rare or late onset, these techniques may not be possible^10^.

Estimating penetrance for a variant of unknown significance identified, for example, during genome sequencing-based screening can be particularly challenging. The problem is exemplified by the large number of reported *SOD1* gene variants in amyotrophic lateral sclerosis (ALS). *SOD1* variants are an important cause of ALS, and over 180 ALS-associated variants in the gene are reported to date^11-13^, but family pedigrees suitable for establishing penetrance are available for only a minority of these.

We have developed a new method to calculate penetrance for variants with an autosomal dominant inheritance pattern using population level data from unrelated people who are and are not affected by the associated phenotype. It can be operated using variant information drawn only from affected populations, stratified according to family history between ‘familial’ and ‘sporadic’ disease presentations. This approach is based on our previously published model of disease which explains how variant penetrance and sibship size determine the presentation or absence of a disease for families in which the variant occurs^14^.

The method is complementary to and fills an important gap left by existing techniques. Using population-scale data, it takes full advantage of the rapidly growing quantity of genetic data that are being generated for a wide range of human disease and therefore it is ideally placed to be a valuable tool in the precision medicine era. Moreover, the capacity to assess penetrance based on the distribution of a variant between samples of unrelated people drawn only from the affected population allows estimates unbiased by kinship-specific effects or ascertainment of unaffected population members.

We have tested the approach in four variant-disease case examples, drawing upon the most common and widely studied autosomal dominant risk variants for each disease: the p.Gly2019Ser variant of the *LRRK2* (OMIM: 609007) gene for PD^2^; variants in the *BMPR2* (OMIM: 600799) gene for heritable pulmonary arterial hypertension (PAH)^15^; and variants in the *SOD1* (OMIM: 147450) and *C9orf72* (OMIM: 614260) genes for ALS^13, 16^.

## Methods

### Model

The disease model our method builds upon^14^ makes the following assumptions: a rare dominant pathogenic variant is necessary but not sufficient for disease to occur, therefore penetrance is not complete and family members who do not harbour the variant are not affected; all individuals harbouring the variant are ascertained; all variants are inherited from exactly one parent, thus there are no people homozygous for the variant or *de novo* variants.

The model calculates three probabilities for a nuclear family where one parent harbours a given variant: that no family members are affected,*P*(*unaffected*); that exactly one member is affected,*P*(*sporadic*); and that more than one member is affected,*P*(*familial*). These probabilities are determined by penetrance, *f*, and sibship size, *N*. In a family with *N* siblings:

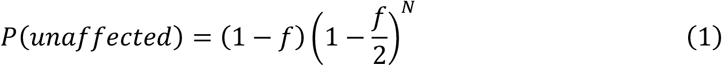

where the parent is unaffected, and none of the sibs are affected (each being transmitted the variant with probability ½).

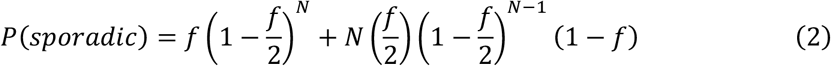

where either the parent is affected and no siblings are affected, or the parent is unaffected and exactly one of the sibs is affected.

Then,

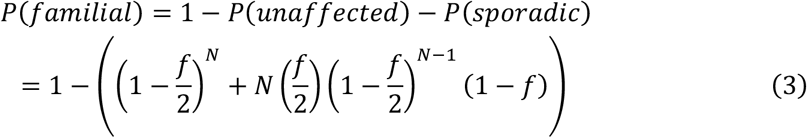

### Application to penetrance calculation

Conversely, given the observed rates of the unaffected, sporadic, and familial disease states in families where the pathogenic variant occurs and the average sibship size for these families, we can estimate penetrance. We can also estimate penetrance based on the observed rates of families presenting as unaffected versus *affected*, a fourth disease state whereby *P*(*affected*) = *P* (*familial*) + *P* (*sporadic*). Observed disease state rates can be specified directly if known for a population or derived as a weighted proportion of estimates of heterozygous variant frequency given across a valid subset of the four defined states (see Table 1); the appropriate weighting factors will vary based on the disease states modelled. Sibship size can be estimated for the sample either directly, based on the average sibship size among the described families, or indirectly, by designating an estimate representative of the sample (e.g. from global databases).

**Table 1.**
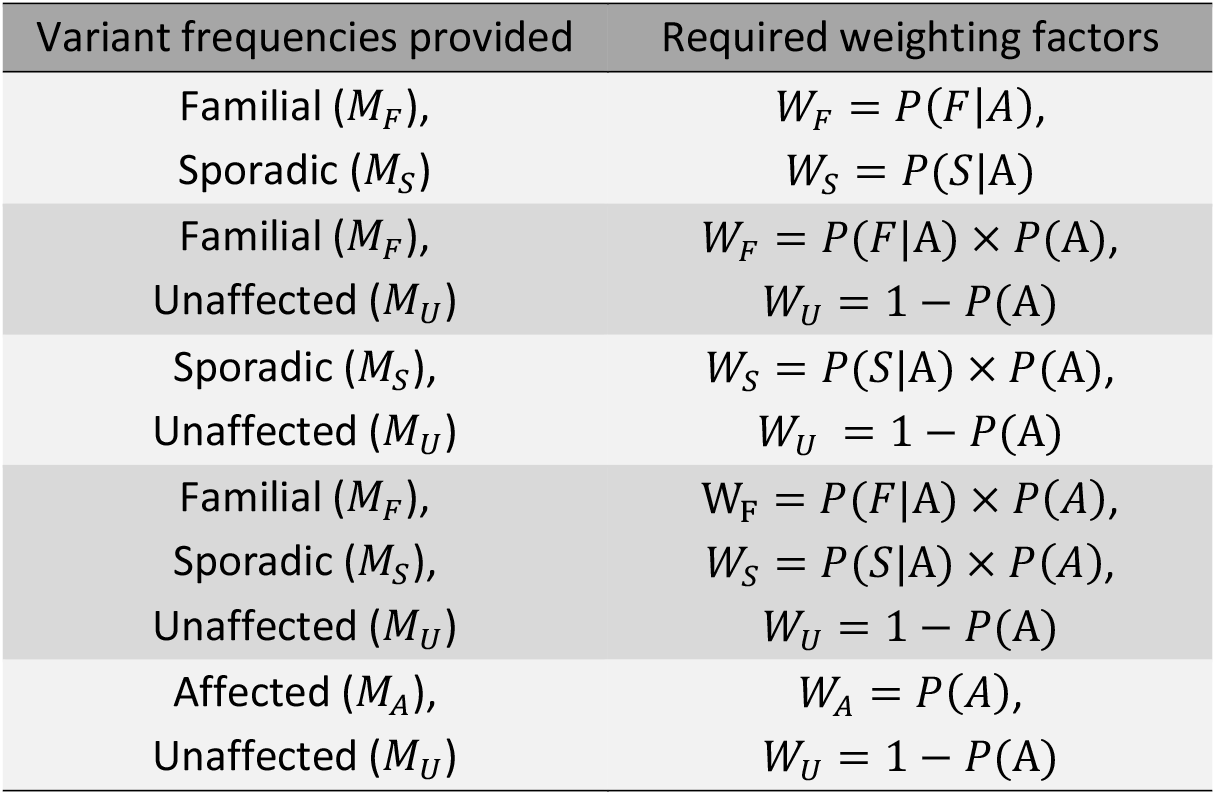
Valid disease state combinations and weighting factors used to estimate disease state rates associated with a given variant as described in Figure 1 and the Supplementary Methods 1.1. M_F,S,U,A_ = variant frequencies in the familial, sporadic, unaffected, and affected states; W_F,S,U,A_ = weighting factors for the familial, sporadic, unaffected, and affected states; P(A) = the probability of a member of the sampled population being affected; P(F|A) = disease familiality rate; P(S|A) = disease sporadic rate.

Our method involves four steps and allows an optional further step for deriving error in the estimate. These processes are summarised in Figure 1 and comprehensively outlined in the Supplementary Methods (1.1-1.2) – including details of simulation studies used to develop and validate the method and comparison between using a lookup table and a maximum-likelihood approach. We assume that: in variant frequency estimates, disease state classifications are stable over time and assigned according to the status of the sampled person and first-degree relatives only; individual families are represented only once in variant frequency estimates; weighting factors and average sibship size represent absolute values; the value specified for sibship size is representative of sibship size across disease state groups; in families harbouring the pathogenic variant, the associated trait can only manifest owing to that variant.

**Figure 1.**
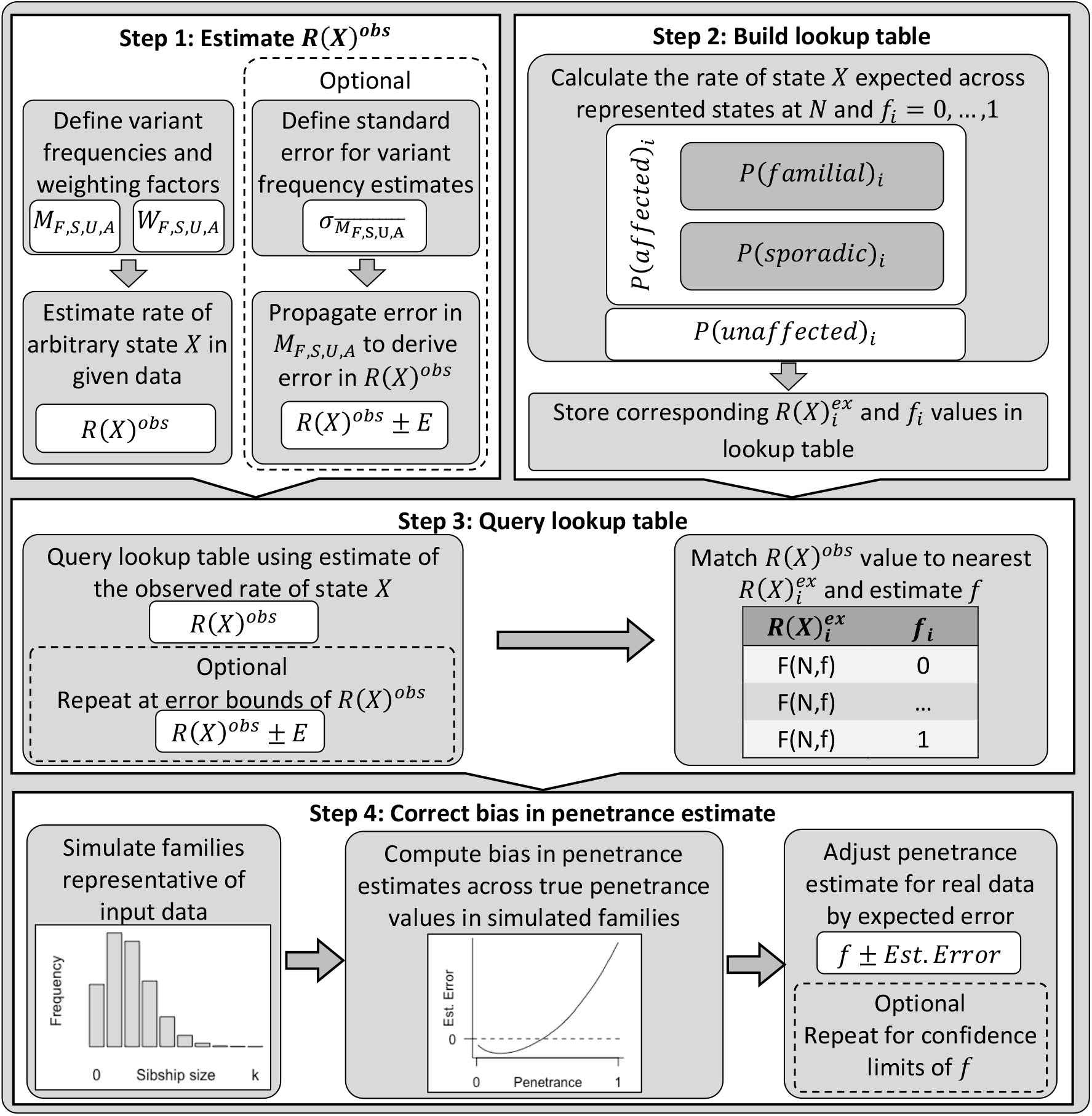
Summary of the key steps within this penetrance estimation approach. Step 1: variant frequencies (M) and weighting factors (W) are defined for a valid subset of the familial (F), sporadic (S), unaffected (U), and affected (A) states (see table 1) to calculate rate of one of these states, arbitrarily labelled state X, among families harbouring the pathogenic variant across those states with data provided, R(X)^obs^. Step 2: Equations 1-3 are applied to calculate P(familial), P(sporadic), P(unaffected), and P(affected), for a series of penetrance values, f_i_ = 0,…, 1, at a defined sibship size, A. The rate of state X expected at each f_i_ among variant harbouring families from those states represented in Step 1,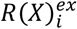, is calculated and stored alongside the corresponding f_i_ in a lookup table. Step 3: The lookup table is queried using R(X)^obs^ to identify the closest 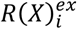 value and corresponding f_i_. Step 4: Bias in the obtained f_i_ estimate is corrected by simulating a population of families representative of the sample data, estimating the difference between true and estimated penetrance values in this population between f = 0,…, 1 and adjusting the estimated f_i_ by error predicted within a polynomial regression model fitted upon the simulated estimate errors. Optional steps: Confidence intervals for R(X)^obs^ can be calculated from error in the estimates of M provided^17^. Penetrance is estimated as in Steps 3 and 4 for the interval bounds. All steps within this approach are comprehensively detailed in Supplementary Methods 1.1.

### Tool access and code availability

This method is available as an R function (R version 3.6.3) and, to facilitate easy use, as a publicly available web resource (https://adpenetrance.rosalind.kcl.ac.uk), developed using the R Shiny package (version 1.4.0.2). The source code of the R library is available on GitHub (https://github.com/ThomasPSpargo/adpenetrance), including the R scripts used for approach validation as described in this manuscript and Supplementary Methods 1.2. The web tool is further described in the Supplementary Methods 1.3 and Figure 2 presents an example of its usage.

**Figure 2.**
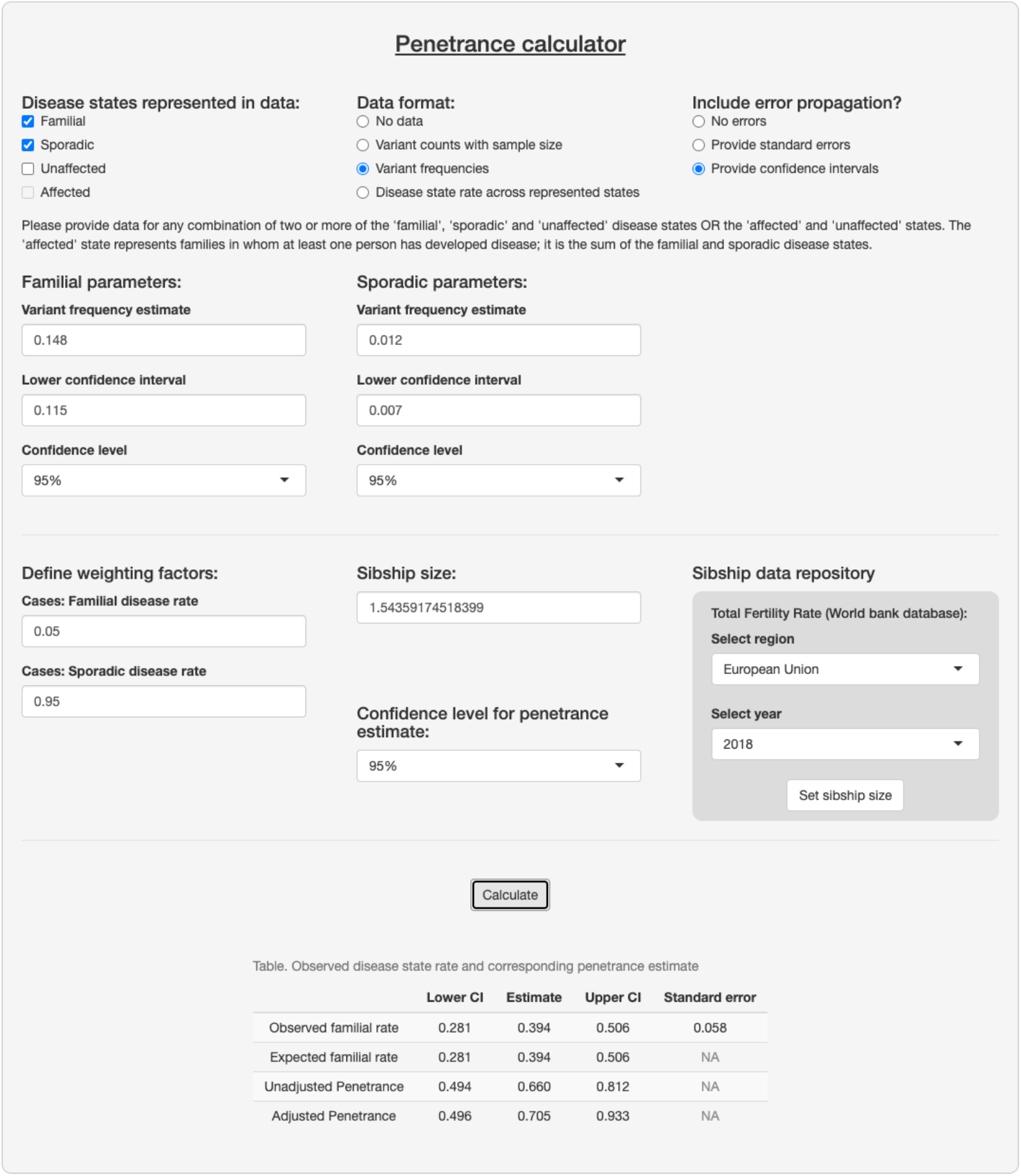
example interface and output of the ADPenetrance web tool (https://adpenetrance.rosalind.kcl.ac.uk). Here we show the example of penetrance of SOD1 variants for amyotrophic lateral sclerosis in a European population, applying variant frequency estimates for familial and sporadic ALS patients of European ancestry and the average Total Fertility Rate for the European Union in 2018^(Refs. 18, 19)^.

### Case examples

Input parameters for included case studies were estimated using publicly available data. We estimated variant frequency in the familial, *M*_*F*_, and sporadic, *M*_*S*_, states in all cases and, in case 1, additionally ascertained this for the unaffected state, *M*_*U*_, from control samples. In all cases, we derived the standard error of these values, 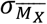, to allow for assessment of error in the penetrance estimate. Variant frequency estimates were weighted to calculate the observed rate of arbitrary state X, *R*(*X*)^*obs*^, among variant-harbouring families from those states modelled using the factors presented in Table 1. Accordingly, the frequencies of familial, *P*(*F*|*A*), and sporadic, *P*(*S*|*A*), disease among the affected population, *A*, were defined in all cases; note that *P*(*S*|*A*)= 1 − *P*(*F*|*A*). The probability of a person being affected, P(A), was defined for case 1 only.

Sibship size, *N*, was estimated in each case based on the Total Fertility Rates reported in the World Bank database^19^ for the world region(s) best representing the sample.

An R script detailing the calculations made for each case study is available within our GitHub repository.

#### Case 1: LRRK2 penetrance for PD

We estimated the penetrance of the p.Gly2019Ser variant of *the LRRK2* gene for PD. This case illustrates the flexibility of this method for application using data drawn from several combinations of the defined disease states.

The first-degree familiality rate of PD, about 0.105, was used to estimate *P*(*F*|*A*) and *P*(*S*|*A*)^20, 21^. *P*(*A*) was estimated as 1 in 37 (0. 027), the lifetime risk of developing PD^22^.

We estimated *M*_*F*.*S*.*U*_ based data aggregated from 24 world populations^23^. Of 5,123 unrelated people with familial PD manifestations, 201 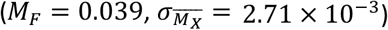 harboured the *LRRK2* p.Gly2019Ser variant, compared to 179 of 14,253 with sporadic PD manifestations 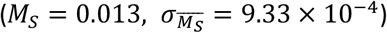 and 11 of 14,886 unaffected controls 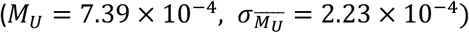 We additionally decomposed this dataset to make region-specific penetrance estimates (see Table S2).

Approximately 75% of the sample is drawn from regions where European ancestry predominates. However, no single region is representative of the total sample. We accordingly estimated that *N* = 1.646 by aggregating Total Fertility Rate estimates available in the World Bank database^19^ across each population sampled, weighted by the proportional contribution of each population to the total sample (see Table S1).

#### Case 2: BMPR2 penetrance for heritable PAH

We estimated the penetrance of variants in the *BMPR2* gene for heritable PAH, a gene for which the low penetrance of pathogenic variants is well established^24^.

Input parameters were defined based on only people with idiopathic (sporadic) or heritable PAH diagnoses^15^. This captures people with and without family disease history and excludes PAH manifestations associated with comorbidities or drug exposure.

We estimated *P*(*F*|*A*)and *P*(*S*|*A*)using the first-degree familiality rate of heritable PAH, about 0.055 of people affected by either idiopathic or familial PAH^24^.

To minimise any study specific bias, we applied data from two reports to build independent estimates for each of *M*_*F,S*_. The first dataset^15^, includes 247 people with familial PAH, of which 202 harboured *BMPR2* variants 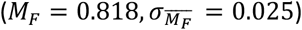, compared to 200 of 1174 in the sporadic state 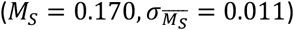 The second dataset^25^ identified that 40 of 58 people with familial PAH 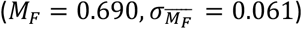 harboured *BMPR2* variants, compared to 26 of 126 in the sporadic state 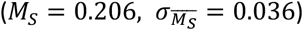 Variant counts are additionally reported separately for small genetic variations (single nucleotide variants and indels) and large genetic rearrangements in *BMPR2*, allowing penetrance estimation stratified by variant type.

The first dataset may violate two assumptions of our approach: first, information on familial clustering was reportedly unavailable and so some families may be represented more than once in the familial state; second, it is not specified whether disease familiality is defined only by the disease status of first-degree relatives. The second sample overcomes a limitation of the first as each family is represented only once in variant counts. However, it is not reported whether disease states are defined according to the status of first-degree relatives only. The correct weighting of *M*_*f,s*_ by the first-degree familial disease rate will minimise any impact of having incorrect familiality definitions applied to the variant frequency estimates upon the final penetrance estimates.

The first cohort samples people from Asian, European, and North American populations; French, German and Italian cohorts comprise about 60% of the sample^15^. The second cohort samples people exclusively from Western Europe^25^. We therefore estimated that *N* = 1.543 in both instances, the Total Fertility Rate of the European Union in 2018^(Ref. 19)^.

#### Cases 3 and 4: SOD1 and C9orf72 penetrance for ALS

We estimated the penetrance of variants in the *SOD1* and *C9orf72* genes for ALS. In *SOD1*, we examined the aggregated penetrance of *SOD1* variants harboured by people with ALS. For *C9orf72*, we examined the penetrance of a single pathogenic variant, a hexanucleotide GGGGCC repeat expansion (*C9orf72*^RE^). These penetrances have been historically difficult to establish without incurring kinship-specific biases. These cases represent ideal candidates for usage of our method.

The first-degree familiality rate of ALS, about 0.050, was applied to define *P*(*F*|*A*) and *P*(*S*|*A*) in these cases^26, 27^.

We drew upon the results of two recent meta-analyses^18, 28^ to estimate *M*_*F,S*_ for *SOD1* and *C9orf72*^RE^. As variant frequencies differed between Asian and European ancestries, we modelled penetrance separately for each group. We derived 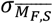 using z-score conversion from the 95% confidence intervals (95% CIs) reported: for the arbitrary state X,

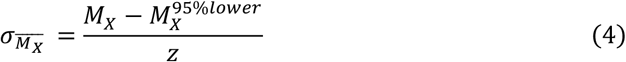

where Z = 1.96 and 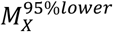 is the lower 95% CI bound of the estimate *M*_*X*_.

In Asian ALS populations: *SOD1* variants were harboured by o.300 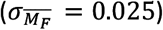 of people with familial and o.o15 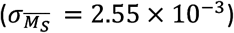 with sporadic disease; *C9orf72*^RE^ was harboured by 0.04 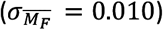 of people with familial and 0.01 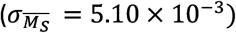 with sporadic disease. In Europeans: *SOD1* variants were harboured by 0.148 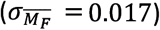 of people with familial and 0.012 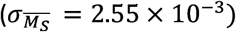 with sporadic disease; *C9orf72*^RE^ was harboured by 0.32 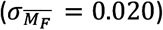 of people with familial and 0.05 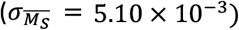 with sporadic disease.

The *SOD1* meta-analysis allowed consideration of the extended kinship when defining familial ALS. The familiality definition used in the *C9orf72* analysis is not stated. The impact of this upon final penetrance estimates should again be minimal, as *M*_*f,s*_ were correctly weighted by the first-degree familiality rate.

In these datasets, the Asian ancestry cohorts were predominantly individuals from East Asia, with small proportion from South Asia. The European ancestry cohorts primarily comprise people from European countries, with some from North America and Australasia. Accordingly, 3 was estimated for the Asian population samples as 1.823, the Total Fertility Rate for East Asia and Pacific in 2018, and for the European population as 1.543, the Total Fertility Rate for the European Union in 2018^(Ref. 19)^.

## Results

Here we summarise the input data and results of the case studies modelled (see Table 2).

**Table 2.**
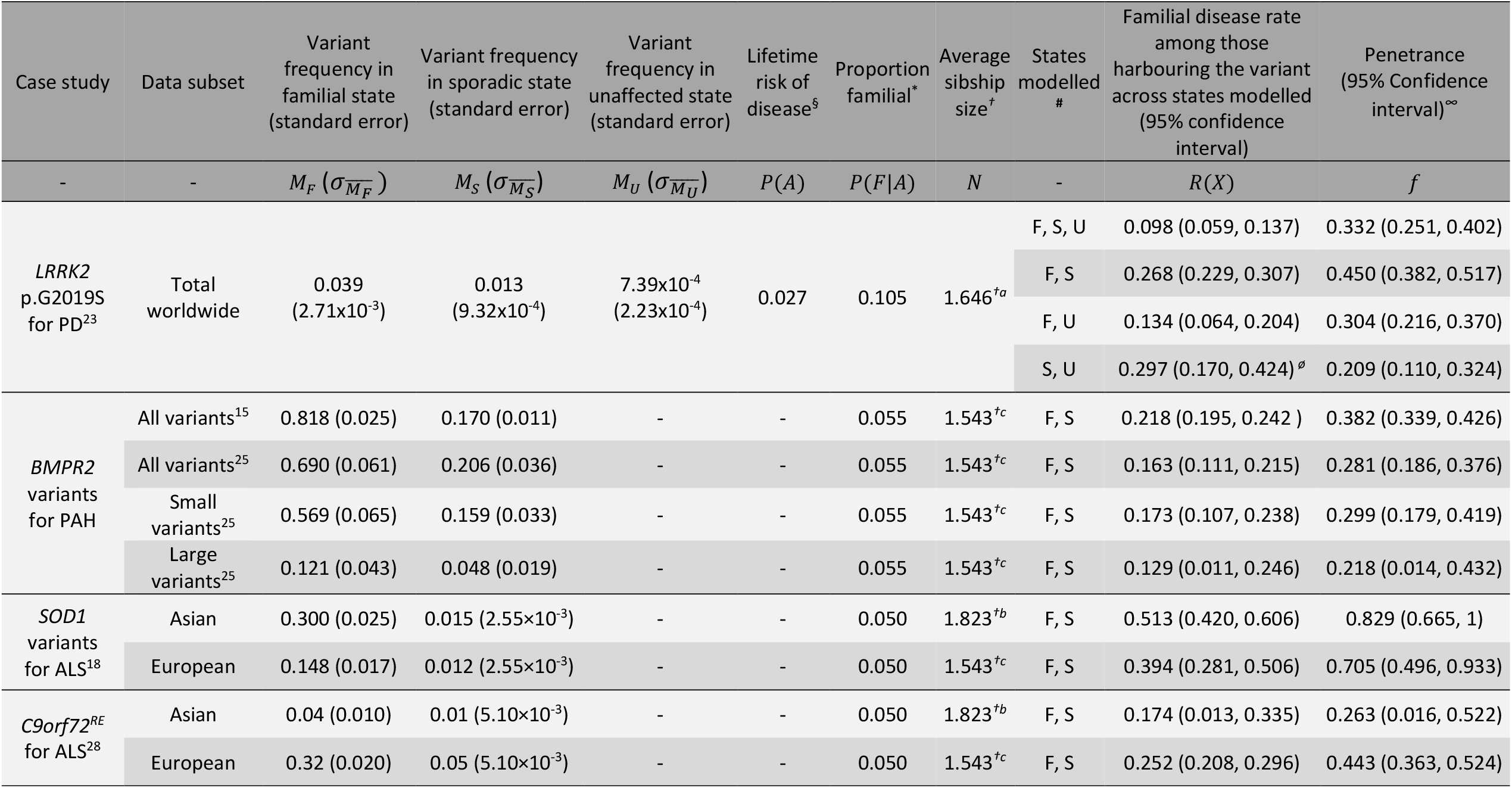
Penetrance estimation for the present case studies. ^§^Lifetime disease risk is only required as a weighting factor where the unaffected (control) population are represented within the data given (see Table 1); ^*^Proportion sporadic is defined as 1 – proportion familial (P(S/A) = 1 − P(F/A)); ^†^Estimated using Total Fertility Rates reported for the: populations sampled to calculate variant frequencies (see Table S1) ^†a^, East Asia and Pacific^†b^, or European Union^†c^ regions in 2018^(Ref. 19)^; ^**#**^F=familial, S=sporadic, U=unaffected (controls); ^ø^Rate of sporadic disease has been calculated here because the familial state is not represented; C9orf72^RE^ = the pathogenic C9orf72 GGGGCC hexanucleotide repeat expansion. ^∞^Step 4 penetrance estimates are shown here, see Table S5 for unadjusted penetrance estimates derived in Step 3.

In case 1, we estimated the penetrance of the *LRRK2* gene p.Gly2019Ser variant for PD. The estimates made (see Table 2) were consistent across the modelled disease state combinations, with some discordance between estimates derived with and without the inclusion of the unaffected disease state. This divergence reflects that the variant is rare in the unaffected (control) population and the accompanying issues of harmonising data from different ancestry groups. Penetrance estimates made across the same four disease state combinations for a European ancestry subsample of this dataset were all between 0.429 (95 %*CI*: 0.348, 0.509) and 0.548 (95 % *CI*: 0.212, 0.951). Penetrance estimates for each sub-population are shown in Table S2.

In case 2, we estimated the penetrance of variants in *BMPR2* for PAH (see Table 2). Slightly higher penetrance was estimated using the first sample set^15^ than in the second^25^ and penetrance was comparable between the defined *BMPR2* variant subtypes of the second sample set. The differences in these estimates reflect variation in *M*_*F,S*_ between the cohorts and may be affected by unspecified family clustering within the first sample set. It is not known for either dataset whether family history classifications were restricted to first-degree relatives only and so the estimates obtained may be slightly inflated. With the available data these possibilities cannot be explored further.

In cases 3 and 4, we estimated the penetrance of variants in *SOD1* and *C9orf72* for ALS (see Table 2). The estimates obtained demonstrate consistency within genes across populations and indicate that the penetrance for ALS is greater in people harbouring *SOD1* variants than in those harbouring *C9orf72*^RE^. Table S3 presents additional penetrance estimates made for widely-described *SOD1* variants: penetrance was estimated as 1 for p.Ala5Val, 0.648 for p.Ile114Thr, and 0.0001 for p.Asp91Ala. Each estimate made in these case studies may be slightly inflated owing to inclusion of extended kinship within familiality definitions.

## Discussion

We have developed a novel approach to estimate the penetrance of genetic variants pathogenic for autosomal dominant traits. The method was tested via simulation studies (see Supplementary Methods 1.2) and application to several case studies.

Our penetrance estimates of the *LRRK2* p.Gly2019Ser variant for PD and of the aggregate penetrance of *BMPR2* variants for PAH closely matched those obtained using other approaches. Previous research on *LRRK2* p.Gly2019Ser estimates its lifetime penetrance between 0.24 (95 % *CI*: 0.135, 0.437) and 0.45 (no CI reported) when analysing data that is not liable to inflation owing to biased selection of familial cases^2^. Longitudinal analysis of disease trends among 53 families harbouring *BMPR2* variants finds penetrance as 0.27 overall, 0.42 for women and 0.14 for men^29^.

The estimates in the *SOD1* and *C9orf72* case studies align with current understanding of penetrance in these ALS genes.

For *SOD1* variants, penetrance for ALS is incomplete and differs between variants^10, 30^. The widely-described p.Ala5Val (formally p.Ala4Val) variant has a recorded penetrance of 0.91 by age 70^(Ref. 31)^. Among other variants, penetrance is typically lower^10, 30^. Of those best characterised, p.Ile114Thr approaches complete penetrance in some pedigrees and p.Asp91Ala reaches polymorphic frequency in some populations, with linked ALS presentations typically displaying an autosomal recessive pattern^10, 11, 31^. Our estimates for heterozygous inheritance of these individual variants aligned with these observations (see Table S3) and highlight the spectrum of penetrance across variants in *SOD1*. Our estimate for the p.Asp91Ala variant in particular supports the hypothesis that it is associated with ALS via a recessive or oligogenic inheritance pattern^32^. The absence of p.Asp91Ala within the familial ALS database sampled further corroborates the finding. Accordingly, our penetrance estimates in Asian and European populations can be taken to suitably represent an aggregated penetrance of risk variants in *SOD1* for ALS; some variation between populations can be expected, reflecting differences in the admix of variants between them.

For *C9orf72*, we modelled the penetrance of its pathogenic hexanucleotide repeat expansion for ALS. Pleiotropy of this variant is widely reported, additionally conferring risk for frontotemporal dementia and, to a lesser degree, other neuropsychiatric conditions^33^. Past penetrance estimates made for this variant are vulnerable to inflation from biased ascertainment of affected people, and the variant is more common among unaffected people than would be expected if these estimates were accurate^16, 33, 34^. A previous study tentatively reports the penetrance of this variant for either ALS or frontotemporal dementia as 0.90 by age 83 after attempting to adjust for ascertainment bias within their sample^34^. Accounting for lifetime risk of each phenotype and their respective familiality rates, people of European ancestry harbouring *C9orf72*^RE^ appear to develop ALS or frontotemporal dementia with comparable frequency, we calculated that 1.012 cases of ALS emerge per case of frontotemporal dementia (See Table S4; ^28, 35-38^). It is therefore reasonable to predict that, if the variant has 0.90 penetrance for the joint condition of ALS and frontotemporal dementia, its penetrance of for ALS alone would be around 0.45. The 0.45 estimate is comparable to the upper bound of our findings. However, we note that our calculation does not account for the common co-occurrence of ALS and frontotemporal dementia and that the true penetrance of *C9orf72*^RE^ for the joint ALS-frontotemporal dementia condition is likely lower than the tentative 0.90 estimate.

The method we present has high validity. Internal validity is demonstrated within simulation studies (see Supplementary Methods 1.2). Criterion and face validity are shown across the present case studies, aligning with estimates made using other techniques and current understanding of the assessed cases. Construct validity is also demonstrated: in the ALS case studies, disease risk was greater for those harbouring a pathogenic *SOD1* variant than for those with the *C9orf92* repeat expansion. This aligns with the multi-step model of ALS^39^, where harbouring *SOD1* variants is associated with a 2-step disease process, converse to the 3-step process associated with *C9orf72*^RE^.

The data necessary to operate the present approach is distinct from that of techniques which examine patterns of disease among affected people, allowing assessment of penetrance in unrelated populations rather than families. The estimates are therefore unaffected by kinship-specific modifiers and are instead applicable to the sampled population.

Where analysis is confined to people affected by disease, across the familial and sporadic states, we circumvent the ascertainment biases affecting designs which examine the distribution of a variant between affected and unaffected populations^9^. Where analysis includes data for unaffected samples (i.e. controls harbouring the variant) these would not be avoided; ascertainment of controls compared to cases has equivalent challenges irrespective of the penetrance estimation approach. However, as our method does not require this information if data of familial and sporadic cases are available, this does not majorly limit the approach.

Furthermore, limitations of ascertainment will diminish as huge datasets of genetic and phenotypic information available within public databases become increasingly available. Therefore, the usefulness of penetrance estimates generated through population data will grow as the size and scope of genetic data held in such datasets expands^9^.

A limitation of this approach is the definition of familiality, which is the occurrence of the studied trait in a first-degree relative under this model. In practice, familial disease may be defined using various criteria, for example considering the disease status of second- or third-degree relatives, or including related diseases that may share a genetic basis^27, 40^. For example, ALS and frontotemporal dementia each share a genetic basis, and considering a family history of frontotemporal dementia is reasonable when assessing familiality in a person with ALS. If the extended kinship is incorporated within familial disease state definitions, then the familial rate will trend upwards and inflate penetrance estimates. Using a wider definition of being affected is acceptable, although it will yield penetrance estimates for the joint condition.

A further caveat is that the model equations assume a particular family structure. It is not feasible to include all possible family configurations for large quantities of summary data however and approximations made are sufficiently close to provide an estimate of penetrance.

This method is suitable for calculating the point, rather than age-dependent, penetrance of pathogenic variants and can be applied to any germline genetic variation associated with a disease via an autosomal dominant inheritance pattern. Penetrance can be derived for an individual variant or for an aggregated set of variants, with the latter indicating an averaged burden of variants meeting the given criteria. We suggest that confidence intervals should be included when using this approach; the size of the interval returned will provide a useful indication of whether the input data is sufficient to accurately estimate penetrance.

The method assumes the stability of disease states among sampled families over time. Such an assumption is typical in case-control research designs, which expect that members of the control sample will not later become cases. However, in traits with age-dependent penetrance, estimates would be influenced by the age of people within sampled families. Younger samples would yield reduced estimates if fewer than two family members are affected when sampled and others will only later become affected. A lifetime penetrance estimate would therefore be most suitably obtained by sampling people beyond the typical age of onset for the studied disease. Age-independent estimates are therefore limited in their clinical utility of indicating disease risk to a person harbouring a tested variant within a given timeframe.

Despite that, point penetrance estimates have several applications, for instance, improving characterisation of pathogenic variants at a population level, facilitating research involving tested variants and, in particular, aiding clinical trial design by supporting the curation of homogenous study populations. They would have additional utility once gene therapies move towards preventative treatment, giving justification for or against such treatment after accounting for possible side effects and risks.

In a scenario where penetrance can be estimated via multiple approaches, we recommend applying each applicable method, given the complimentary nature of these techniques. If the results of multiple approaches conflict, we would suggest inspection of the suitability of the input data given for each method and to prioritise the result which these best fit.

In conclusion, our novel method for penetrance estimation fills an important gap in medical genetics because, making use of the available amounts of population-scale data, it enables the unbiased and valid calculation of penetrance in genetic disease instances that would be otherwise difficult or impossible using existing methods. It serves to expand the range of genetic diseases and variants for which high-quality penetrance estimates can be obtained, as we illustrate in the ALS case examples. Estimates drawn via this approach have clear utility and will be useful for characterisation of pathogenic variants, with benefits for both clinical practice and research. They have wider relevance to the population than those obtained by studying particular kinships and will be more interpretable for clinical professionals.

The tool code is available on GitHub (https://github.com/ThomasPSpargo/adpenetrance) and the method is available and free to use via a public webserver (https://adpenetrance.rosalind.kcl.ac.uk).

## Data Availability

All data referred to in the manuscript is publicly available the original sources are referenced within.
An R library containing the relevant scripts for our method, including validation and analysis of case studies, is hosted on GitHub: https://github.com/ThomasPSpargo/adpenetrance

## Conflict of interest statement

CML sits on the SAB for Myriad Neuroscience. AAC reports consultancies or advisory boards for Amylyx, Apellis, Biogen Idec, Brainstorm, Cytokinetics, GSK, Lilly, Mitsubishi Tanabe Pharma, Novartis, OrionPharma, Quralis, and Wave Pharmaceuticals

## Acknowledgements

This is an EU Joint Programme-Neurodegenerative Disease Research (JPND) project. The project is supported through the following funding organizations under the aegis of JPND-http://www.neurodegenerationresearch.eu/ (United Kingdom, Medical Research Council MR/L501529/1 to A.A.-C., principal investigator [PI] and MR/R024804/1 to A.A.-C., PI]; Economic and Social Research Council ES/L008238/1 to A.A.-C. [co-PI]) and through the Motor Neurone Disease Association. This study represents independent research partly funded by the National Institute for Health Research (NIHR) Biomedical Research Centre at South London and Maudsley NHS Foundation Trust and King’s College London. The work leading up to this publication was funded by the European Community’s Horizon 2020 Programme (H2020-PHC-2014-two-stage; grant 633413). We acknowledge use of the research computing facility at King’s College London, Rosalind (https://rosalind.kcl.ac.uk), which is delivered in partnership with the National Institute for Health Research (NIHR) Biomedical Research Centres at South London & Maudsley and Guy’s & St. Thomas’ NHS Foundation Trusts and part-funded by capital equipment grants from the Maudsley Charity (award 980) and Guy’s and St Thomas’ Charity (TR130505). The views expressed are those of the author(s) and not necessarily those of the NHS, the NIHR, King’s College London, or the Department of Health and Social Care.

## Notes

### Author Declarations

This work involved analysis of already published data only and as such IRB approval was not required.

### Summary of Updates

Updated penetrance estimation approach; additional steps taken to validate approach (presented in Supplementary Methods); reanalysis of all case studies with revised method; additional analyses for sub-populations included within Case Study 1 (Table S2); revised discussion to address further considerations relevant to the method and penetrance estimates made.

