## Supplementary Materials for "Calculating variant penetrance using family history of disease and population data Authorship"


1. Supplementary Methods
   1.1. Penetrance calculation procedure
   1.2. Approach Validation and testing
      1.2.1. Lookup table validation: an alternative maximum-likelihood approach
      1.2.2. Simulation studies
   1.3. ADPenetrance: a companion web tool

2. Supplementary Tables
   2.1. Table S1 – Sample characteristics and calculation of N for data applied in case study 1
   2.2. Table S2 – Penetrance estimation of the *LRRK2* p.Gly2019Ser variant for Parkinson’s Disease across populations sampled in case study 1
   2.3. Table S3 – Penetrance estimation for widely described *SOD1* variants
   2.4. Table S4 – Estimation of the incidence of amyotrophic lateral sclerosis relative to frontotemporal dementia among people of European ancestry who harbour the pathogenic hexanucleotide GGGGCC repeat expansion of the *C9orf72* gene
   2.5. Table S5 - Comparison of unadjusted penetrance estimates derived for the case studies presented in Table 2 between the lookup table and maximum-likelihood approaches

### 1. Supplementary Methods

#### *1.1. Penetrance calculation procedure*

Here follows an outline of the present approach to penetrance estimation. This method is available as an R function (R Version 3.6.2) accessible at https://github.com/ThomasPSpargo/adpenetrance/.

Step 1:
To calculate penetrance using this method, we must identify the rate at which one of the defined disease states (familial, sporadic, unaffected, affected) occurs in families where the variant occurs sampled across a valid combination of two or three of these states. This rate is denoted as $R(X)$, and X can be any one of the four disease states, providing that variant characteristics are reported for that state.

Definitions:
Familial = more than one family member affected
Sporadic = only one family member affected
Unaffected = no family member affected
Affected = at least one family member affected – familial or sporadic not specified.

In Step 1, we determine $R(X)$ as it is observed within input data, $R(X)^{obs}$. If known, $R(X)^{obs}$ can be specified directly, alongside a corresponding indication of the states from which this estimate is derived. If the familial state is represented within input data, then state X is familial. If only the sporadic and unaffected states are represented, then state X is sporadic. If the affected and unaffected states are represented, then state X is affected.

$R(X)^{obs}$ can also be derived as a weighted proportion of heterozygous variant frequency estimates drawn from samples of unrelated people from two or three of the familial, sporadic, and unaffected disease states or the affected and unaffected states. When variant frequency estimates for the familial or sporadic states are included, the frequency of familial, $P(F|A)$, and sporadic, $P(S|A)$, disease among the affected population, A, must feature in weightings; note that, as familial and sporadic states are binary outcomes within the affected population, $P(S|A) = 1 - P(F|A)$. Where the unaffected or affected groups are represented, baseline (e.g. lifetime) risk of a population member being affected, $P(A)$, must be included within weightings.

In this weighted proportion calculation, we respectively denote variant frequencies for familial, sporadic, unaffected, and affected states as $M_{F,S,U,A}$, to be weighted by the factors $W_{F,S,U,A}$. Given that representation of any two or three of the familial, sporadic, and unaffected disease states or the affected and unaffected states can be used estimate $R(X)^{obs}$, we let the familial, sporadic, unaffected, and affected states be arbitrarily denoted as the states $X$, $Y$, and $Z$. Accordingly, letting $M_{F,S,U,A}$ and $W_{F,S,U,A}$ arbitrarily be $M_{X,Y,Z}$ and $W_{X,Y,Z}$ for the states $X$, $Y$ and $Z$,

$$R(X)^{obs} = \frac{M_X W_X}{M_X W_X + M_Y W_Y} \qquad \text{(S1)}$$

if data are given for a valid combination of two disease states, or

$$R(X)^{obs} = \frac{M_X W_X}{M_X W_X + M_Y W_Y + M_Z W_z} \tag{S2}$$

if data are given for the familial, sporadic, and unaffected disease states. Note that all 4 states cannot be specified together as the familial and sporadic states are subsumed within the affected state. For this reason, it is also unsuitable to represent the affected state alongside data for either or both of the familial or sporadic states. Table 1 presents all possible disease state combinations and outlines how the associated weighting factors should be defined to calculate $R(X)^{obs}$.

Step 2:
A lookup table to which $R(X)^{obs}$ can be compared for penetrance estimation is generated here. This table stores a series of $R(X)$ values that would be expected at a given value of penetrance, $f_i$, in a population with average sibship size $N$. We denote this series of $R(X)$ values as $R(X)_i^{ex}$. The sibship size $N$ must be defined alongside the data provided for Step 1 and should represent the average sibship size of the sample from which $R(X)^{obs}$ is determined.

$P(familial)$, $P(sporadic)$, and $P(unaffected)$ are first calculated, following equations 1-3, for a sequence of $f$ values, $f_i = (0.0000, 0.0001, \ldots, 1.0000)$, at a specified $N$. This produces a series of values for each disease state: $P(familial)_i$, $P(sporadic)_i$, and $P(unaffected)_i$. If the affected state has been specified in Step 1, we calculate the probability of being affected, $P(affected)$, at each $f_i$: $P(affected)_i = P(familial)_i + P(sporadic)_i$. $R(X)_i^{ex}$ can then be derived as an unweighted proportion from $P(familial)_i$, $P(sporadic)_i$, $P(unaffected)_i$, and $P(affected)_i$, taking those disease states which were previously represented in Step 1 and ensuring $X$ represents the same state as before. The lookup table is next constructed, storing an index of corresponding $R(X)_i^{ex}$ and $f_i$ values.

Step 3:
The lookup table generated in Step 2 is queried using the $R(X)^{obs}$ estimate obtained in Step 1. The value of $R(X)_i^{ex}$ closest to $R(X)^{obs}$ is identified and the corresponding penetrance value is taken (See Supplementary Methods 1.2.1 for comparison to a maximum-likelihood approach). This value is an uncorrected penetrance estimate, $f^{unadjusted}$, subject to a systematic bias within the approach and should not therefore be taken as the final estimate of penetrance to be determined in step 4. Note that $R(X)^{obs} \approx R(X)_i^{ex}$ unless $R(X)^{obs}$ exceeds or is less than the rate of state $X$ expected between $f = 0, \ldots, 1$ at $N$.

Step 4:
This step computes the final penetrance estimate to be returned by the method, $f^{adjusted}$. It corrects for systematic bias in the $f^{unadjusted}$ estimate from in Step 3, which diverges from the true penetrance value according to the combination of states modelled, the value of penetrance, and the structure of families sampled (see Figures S1, S2).

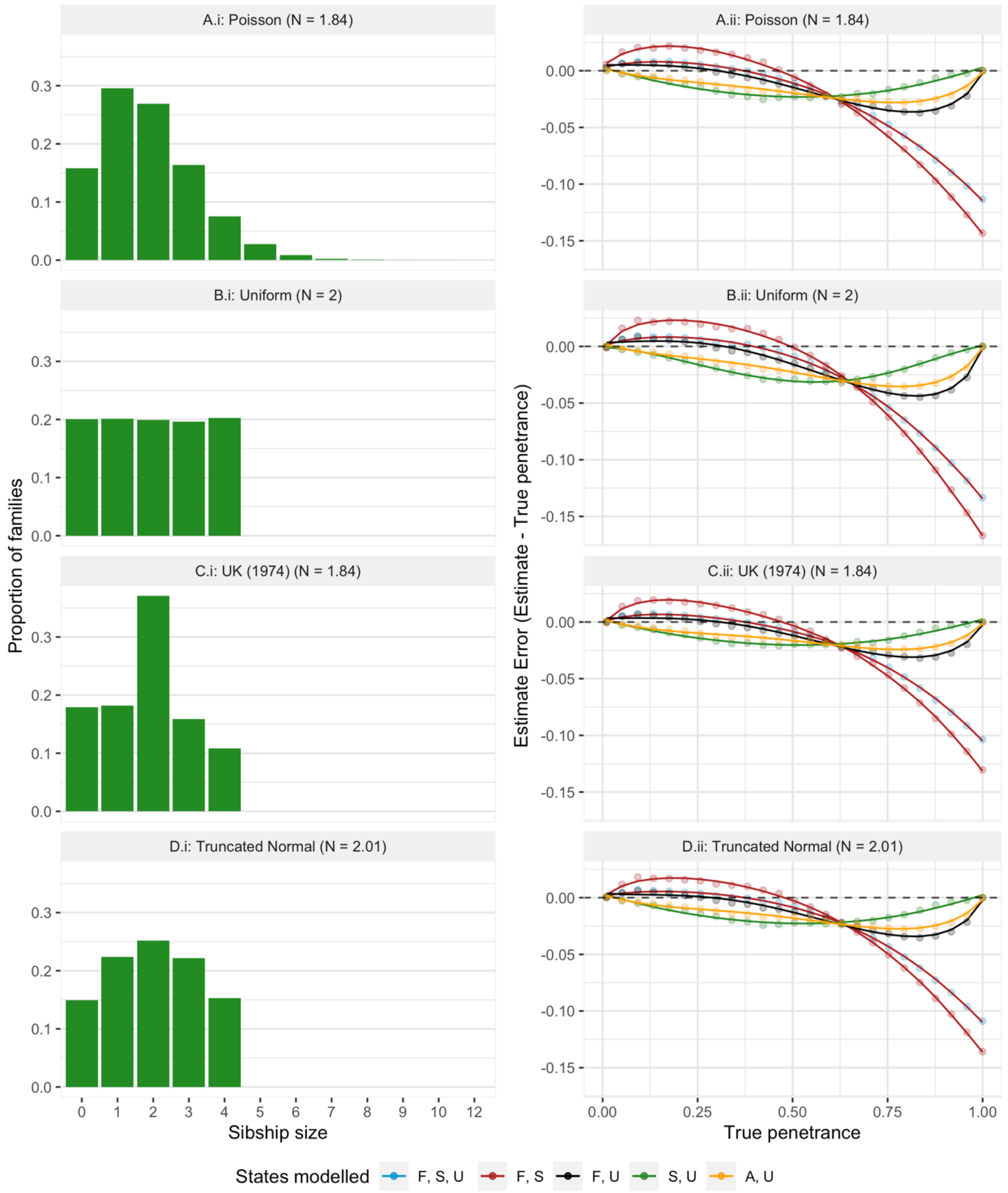


*Figure S1. Errors in unadjusted penetrance estimates across true penetrance values and according to states modelled for a simulated population where sibship sizes follow a given distribution. N = mean sibship size, F = familial, S = sporadic, U = unaffected, A = affected. Panels A.i-D.i show the distribution of sibship sizes across simulated families. Panels A.ii-D.ii display errors in penetrance estimates associated with the corresponding population structure - zero indicates a perfect penetrance estimate, positive values indicate overestimation and negative values underestimation; plotted points display raw error values calculated at each true penetrance value and plotted lines display error values predicted under a fitted polynomial regression model.*

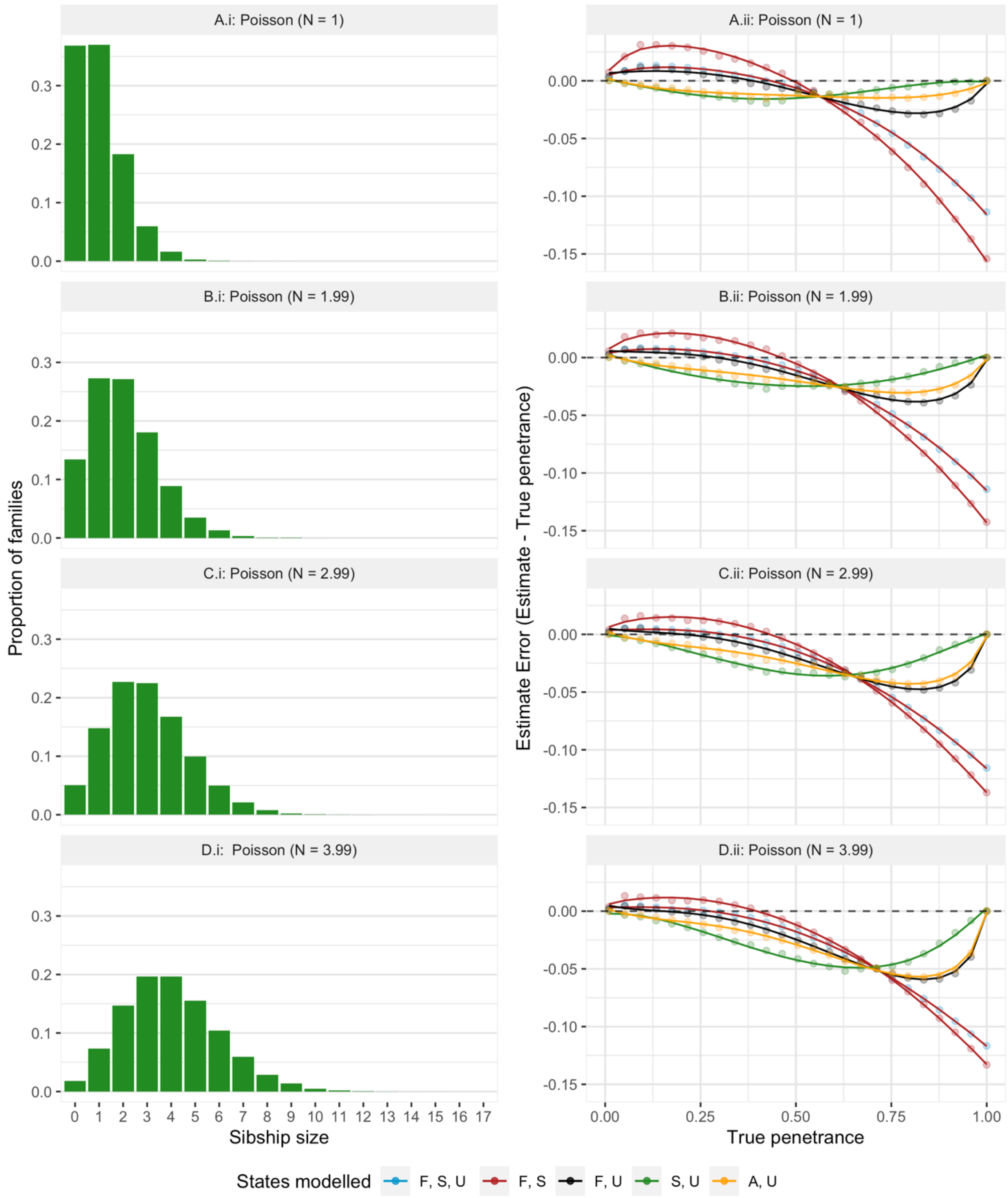


*Figure S2. Errors in unadjusted penetrance estimates across true penetrance values and according to states modelled for a simulated population where sibship sizes follow a Poisson distribution varying by mean sibship size (lambda). N = mean sibship size, F = familial, S = sporadic, U = unaffected, A = affected. Panels A.i-D.i show the distribution of sibship sizes across simulated families. Panels A.ii-D.ii display errors in penetrance estimates associated with each corresponding population structure - zero indicates a perfect penetrance estimate, positive values indicate overestimation and negative values underestimation; plotted points display raw error values calculated at each true penetrance value and plotted lines display error values predicted under a fitted polynomial regression model.*

In Step 4, firstly, a simulated dataset of 90,000 families is pseudo-randomly generated, where each simulated family is assigned a sibship size of the value $N_i^{(sim)}$. The population generated in this step aims to approximate the sibship structure of the real population sampled for penetrance estimation. To ensure replicability, all pseudo-randomisation in this step is performed using the R seed 24.

By default, simulated sibships follow a Poisson distribution with the lambda defined by the mean sibship size, $N$, specified for the real sample data. Example simulated Poisson sibship distributions are presented in Figure S2 A.i-D.i. The Poisson distribution was selected as the default simulation distribution as it is a discrete probability distribution useful for estimating the number of events expected to occur within a given time frame. In this instance, an event is having a child (1 sib) and the time frame is the childbearing years for that family. We note that the Poisson distribution assumes the independence of events and that this assumption would not hold in the present instance (i.e. in real populations, the probability of having additional offspring will be influenced by having already had $N_i$ offspring). However, Figure S1 demonstrates that the degree of error made in Step 3 penetrance estimates is comparable between the Poisson distribution (Panel A.ii) and other hypothetical population structures(Panels B.ii-D.ii), including the distribution shown in C.i, which resembles that of a UK 1974 population birth cohort[1]. Therefore, a simulated population in which sib-sizes follow a Poisson distribution can be considered sufficient for approximating the expected error in unadjusted penetrance estimates made using data from randomly sampled populations. This is corroborated by the results of the simulations presented in supplementary methods (1.2.2).

If the structure of sibships in the real sample is known, then the user can optionally supply the R function with either a vector containing all the sampled sibship sizes or a summary of the sibship distribution, declaring the sibship sizes contained in the sample and the proportion of the sample each sib-size represents. When sibship data are supplied, a simulated sibship distribution is generated based on these data, including only the sibship sizes represented and following its sibship distribution. This tailored simulation population will give more precise $f^{adjusted}$ estimates than those obtained using the default Poisson distribution (see supplementary methods 1.2.2). However, the Poisson distribution is sufficiently precise for adjustment when the sibship distribution of the real data are unknown, under the assumption that population sampling is random and does not exclude families of a particular sibship size (e.g. families of sibship size 0 are not excluded).

A sequence of 25 penetrance values between 0.01 and 1 is also defined, representing true penetrance values of a simulated variant, $f^{true\ (sim)}$. For each $f_i^{true\ (sim)}$, equations 1-3 are applied at each of the $N_i^{(sim)}$ sibship sizes within the simulated population to determine the probability of a family of sibship size $N_i^{sim}$ being familial, sporadic, or unaffected at that $f_i^{true\ (sim)}$. One of the familial, sporadic, and unaffected states is pseudo-randomly assigned to each simulated family, according to the probabilities expected at their sibship size and the given $f_i^{true\ (sim)}$. An unadjusted Penetrance estimate is then made for the simulated population, $f_i^{unadjusted\ (sim)}$, according to the mean sibship size of the simulated population $N^{(sim)}$, and the $R(X)$ observed, $R(X)^{obs\ (sim)}$. Here, $N^{(sim)} \approx N$, with small variation between these values reflecting the pseudo-randomisation of population generation, and State X is defined as in Step 1, with $R(X)^{obs\ (sim)}$ being calculated as an unweighted proportion of the probabilities of X across the modelled states within the simulated dataset.

The difference between corresponding values of $f_i^{unadjusted\ (sim)}$ and $f_i^{true\ (sim)}$ is calculated: $f_i^{error\ (sim)} = f_i^{true\ (sim)} - f_i^{unadjusted\ (sim)}$. A positive $f_i^{error\ (sim)}$ indicates underestimation of penetrance, while negative values denote overestimation. The relationship between $f^{error\ (sim)}$ and $f^{unadjusted\ (sim)}$ is then established by fitting an nth degree polynomial regression model, which, by extension, also indicates the relationship between $f^{error\ (sim)}$ and $f^{true\ (sim)}$. Polynomial models between 1 and 5 degrees are tested, and the best fitting model is selected based on the Akaike Information Criterion. Figures S1 (A.ii) and S2 (A.ii-D.ii) display examples of these error curves fitted for simulated populations where sibship sizes follow a Poisson distribution. Dynamic generation of these regression models is necessary to account for required changes in model fit according to population sibship structure (see Figure S2)

The fitted polynomial regression model is then used to predict error in the penetrance estimate made for the real dataset in Step 3 based on the value of $f_i^{unadjusted}$, $f_i^{error\ (predicted)}$. The final penetrance estimate is then determined: $f_i^{adjusted} = f_i^{unadjusted} + f_i^{error\ (predicted)}$. The validity of these penetrance estimates is demonstrated in the simulation studies presented in Supplementary Methods 1.2.2).

Optional step:
Confidence intervals for the penetrance estimate can be derived through the calculus approach to error propagation[2]. For this, standard errors, $\sigma_{\overline{M_{X,Y,Z}}}$, of the variant frequency estimates given in Step 1 are required. Using these errors, we calculate the standard error in of $R(X)^{obs}$, $\sigma_{\overline{R(X)^{obs}}}$:

$$\sigma_{\overline{R(X)^{obs}}} = \sqrt{\left(\frac{\partial R(X)^{obs}}{\partial M_X}\right)^2 \cdot \sigma_{\overline{M_X}}^2 + \left(\frac{\partial R(X)^{obs}}{\partial M_Y}\right)^2 \cdot \sigma_{\overline{M_Y}}^2 + \cdots} \qquad \text{(S3)}$$

Confidence intervals for $R(X)^{obs}$, $\mathrm{CI}_{\mathrm{R(X)}^{obs}}$, can then be obtained through z-score conversion ($CI_{R(X)^{obs}} = R(X)^{obs} \pm z \times \sigma_{\overline{R(X)^{obs}}}$). The lookup table is then queried as in operation 3 for upper and lower bounds of $\mathrm{CI}_{\mathrm{R(X)}^{obs}}$ to attain upper and lower bounds for the $f_i^{unadjusted}$ estimate obtained in Step 3. These values are then adjusted as in Step 4 according to the fitted polynomial regression model, giving the final penetrance estimates at the confidence interval bounds.

##### 1.2. *Approach validation and testing*

The R scripts used for approach validation are available within our GitHub repository: https://github.com/ThomasPSpargo/adpenetrance/.

###### 1.2.1. *Lookup table validation: an alternative maximum-likelihood approach*

The unadjusted penetrance estimates obtained in Step 3, $f^{unadjusted}$, can also be derived following a maximum likelihood approach. To validate the lookup table approach followed within our method, we additionally derived $f^{unadjusted}$ estimates using Non-Linear Minimisation, leveraging nlm and dbinom functions available within the R stats package (version 3.6.3).

We constructed this validation approach by defining a negative likelihood function which determines, under a binomial distribution, the likelihood of the specified $R(X)^{obs}$ at a given $f^{unadjusted}$ and $N$. Within this function, values of $R(X)^{obs}$ are transformed into integers so that they represent a number of state X events across a certain number of trials (e.g. the rate 0.394 would be multiplied by three orders of magnitude, giving 394 events across 1000 trials). The probability function is defined using equations 1-3, and according to the states modelled in calculating $R(X)^{obs}$.

Non-Linear Minimisation was then applied to determine the most likely $f^{unadjusted}$ given $R(X)^{obs}$ and $N$. The starting value for minimisation was defined as the $f^{unadjusted}$ estimate previously determined via the Step 3 lookup approach.

This approach was applied to each of the case studies presented in Table 2 and we found negligible difference between the $f^{unadjusted}$ estimates generated within non-linear minimisation and via the lookup table method (See Table S5). Thus, these findings confirm the validity of the lookup table approach. The alternative maximum-likelihood method was not adopted for penetrance calculation to avoid potential issues in model convergence if starting values are not appropriately defined.

###### 1.2.2. *Simulation studies*

Here we present the results of simulation studies conducted to test the validity of the 4-step approach outlined in Supplementary Methods 1.1. Example of the systematic bias present in penetrance estimates made in Step 3 are given in Figures S1 and S2.

In each study, the performance of the method was tested on simulated populations containing 90,000 families, pseudo-randomly generated based on sibship distributions previously reported in two distinct samples (see figure S3). The simulated datasets used within these studies were generated pseudo-randomly in R with no set seed number.

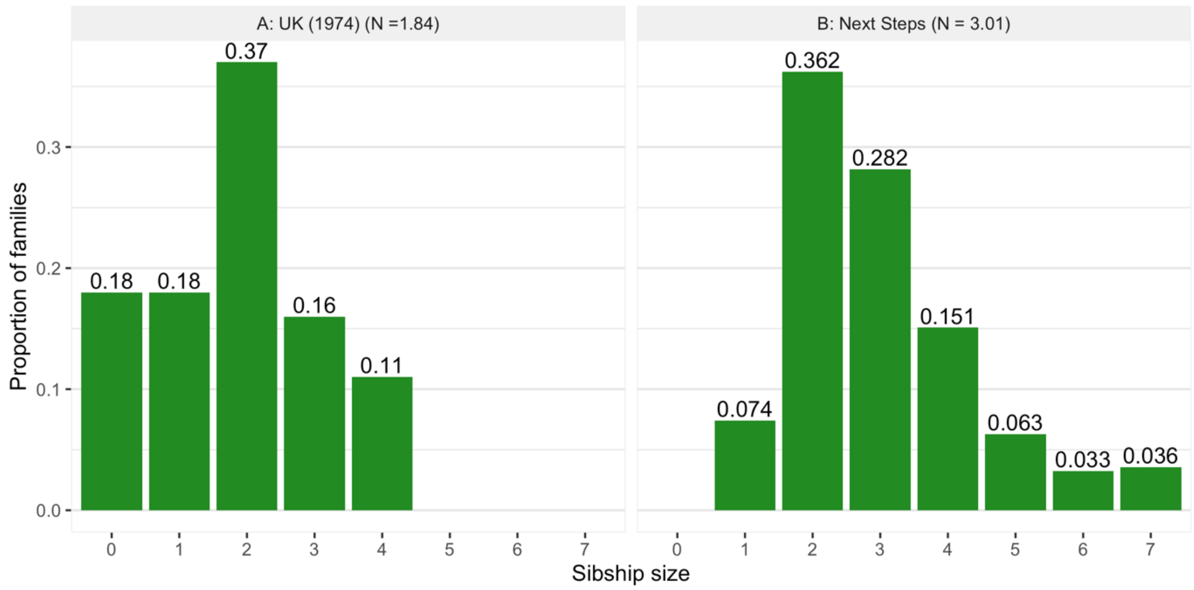


Figure S3. *Sibship distributions upon which simulated populations were modelled across simulation studies. N = mean sibship size. Panel A presents the sibship distribution for the UK population 1974 birth cohort at the completion of their childbearing years; note that the original data reports sibships above size 4 within a collapsed '4 or more' category*[1]*. Panel B presents the sibship distribution across English families sampled in the Next Steps cohort study; note that the original data reports sibships above size 7 within a collapsed '7 or more' category*[3]*.*

The first simulated population (henceforth: the UK population) resembles the sibship distribution across the UK population 1974 birth cohort at the end of their childbearing years (defined as age 45)[1]. The families within this simulated dataset were each pseudo-randomly assigned a sibship size between 0 and 4 according to the probabilities observed in this cohort (See Figure S3) and the mean sibship size, $N$, is 1.84. The simulation population was modelled on these data because they describe the most recent birth cohort for which data is available at the completion of childbearing years and because the distribution is representative of a randomly sampled population. The distribution of sibship sizes across this cohort is comparable to other reported UK and USA birth cohorts[1, 4].

The second population (henceforth: the NS population) was simulated based on the distribution of sibship sizes reported for the Next Steps dataset, a longitudinal sample of children from England[3]. Simulated families were pseudorandomly assigned a sibship size between 1 and 7 according to the probabilities observed in the Next Steps sample (see figure S3) and $N = 3.006$. The simulation cohort was modelled on these data to illustrate the application of the method to a sample which is not fully representative of the population (In this case, the sample does not include families of sibship size 0).

A series of ground truth penetrance values, $f_i^{true}$, were generated for testing within each study. For each $f_i^{true}$, the two simulated populations were generated as described above and the familial, sporadic, and unaffected disease state probabilities expected at each sibship size occuring within a given population were calculated using equations 1-3. One of these three disease states was then pseudo-randomly assigned to each family with the probabilities expected according to the sibship size of that family. Penetrance estimates,

$f_i^{adjusted}$, were then made for the population simulated under the specifications of that study. $f_i^{adjusted}$ estimates were produced for each possible disease state combination, producing in five $f_i^{adjusted}$ estimates for each value of $f_i^{true}$, across the combinations of states modelled. The error in $f_i^{adjusted}$was then determined: $f_i^{error} = f_i^{adjusted} - f_i^{true}$. Positive $f_i^{error}$ values indicate overestimation of penetrance, while negative values indicate underestimation.

In each study, to test the two estimate adjustment approaches allowed in Step 4, we estimated $f_i^{error}$ firstly when the method is supplied no information about the distribution of sibships in the sample data and secondly when this information is supplied. As described in Step 4 (see Supplementary Methods 1.1), the former condition adjusts $f_i^{unadjusted}$ by predicted error in the estimate under a polynomial regression model fitted to a population simulated within the method in which sibships follow a Poisson distribution. The latter condition tailors adjustment of $f_i^{unadjusted}$, by fitting the regression model to a population simulated within the method which approximates the real sample data.

*Validation under correct parameter specification*

We first tested the approach by examining the accuracy of penetrance estimates made using correctly specified input parameters in simulated UK and NS populations harbouring hypothetical variants with known true penetrance values. A sequence of 20 ground truth penetrance values was first defined: $f_i^{true} = (0.05, 0.10, \ldots, 1)$ and the populations were simulated as described above. Penetrance estimates, $f_i^{adjusted}$, were made for these populations, defining $N$ according to the mean sibship size of that sample, approximately 1.84 for the UK and 3.01 for the NS populations, and with $R(X)^{obs}$ calculated across all possible disease state combinations. $f_i^{error}$ was then determined. This simulation was repeated 5 times for each value of $f_i^{true}$, and the results are shown in Figure S4, averaged across repetitions to determine the mean $f_i^{error}$observed at each value of $f_i^{true}$, across each of the disease state combinations. These findings evidence the validity and accuracy of penetrance estimates generated via this approach. They also demonstrate the benefit of supplying information to the function about the distribution of sibships in the sample data when this is known; this benefit is greater if sample data does not accurately represent sibship sizes across the population (e.g. where there are no families of sibship size 0 in the NS dataset).

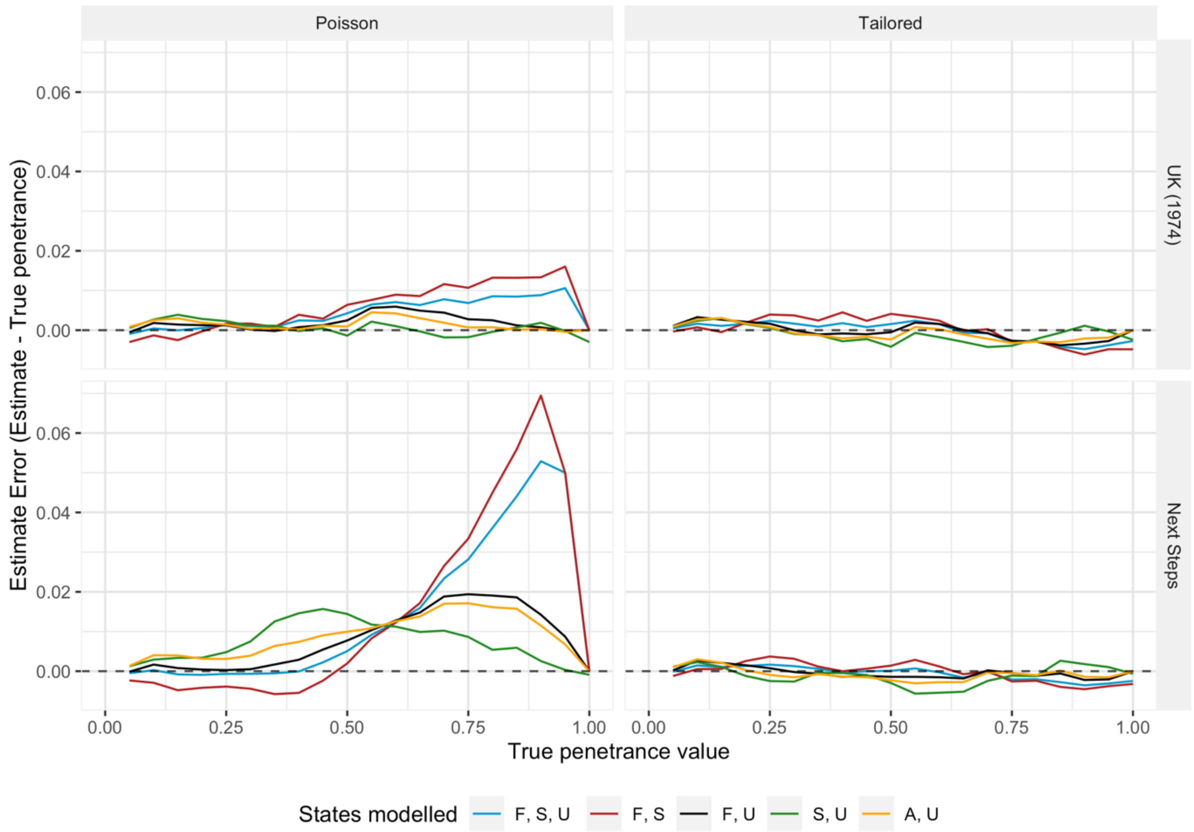


*Figure S4. Error in penetrance estimates across true penetrance values when $R(X)^{obs}$ and $N$ are specified correctly in the simulated UK (1974) and Next Steps populations. Zero indicates a perfect penetrance estimate, positive values indicate overestimation and negative values underestimation. Plot lines represent estimates made when $R(X)^{obs}$ is defined according to different disease state combinations; F = familial, S = sporadic, U = unaffected, and A = affected - state X is represented by the first state named. The panel rows stratify by population simulated (see Figure S3) and columns stratify by Step 4 adjustment approach (see Supplementary Methods 1.1), which follows either the default approach (denoted Poisson) or is tailored to errors predicted under an internally-simulated sibship distribution directly approximating the sample data.*

*Simulation under incorrect parameter specification*

Misspecification of sibship size:

This simulation study examines the accuracy of penetrance estimates when the mean sibship size of sample populations is incorrectly defined. Several values of true penetrance were defined: $f_i^{true} = (0.10, 0.25, 0.50, 0.75, 1.00)$. A sequence of values to represent the degree of misspecification in mean sibship size was also specified: $N_i^{modify} = (-1.5, -1.0, \ldots, 3.0)$. The simulated UK and NS populations were generated as before and estimates of $f_i^{adjusted}$ were made, calculating $R(X)^{obs}$ across all possible disease state combinations and defining $N$ according to the mean sibship size of that sample, approximately 1.84 for the UK and 3.01 for the NS populations, adjusted by each value of $N_i^{modify}$. For instance, if $N = 1.84$ and $N_i^{modify} = -1.5$, penetrance would be estimated based on $N = 0.34$. $f_i^{error}$ was then determined. This simulation was repeated 3 times for each value of $f_i^{true}$, and the results were averaged across these repetitions. These results are presented in Figure S5.

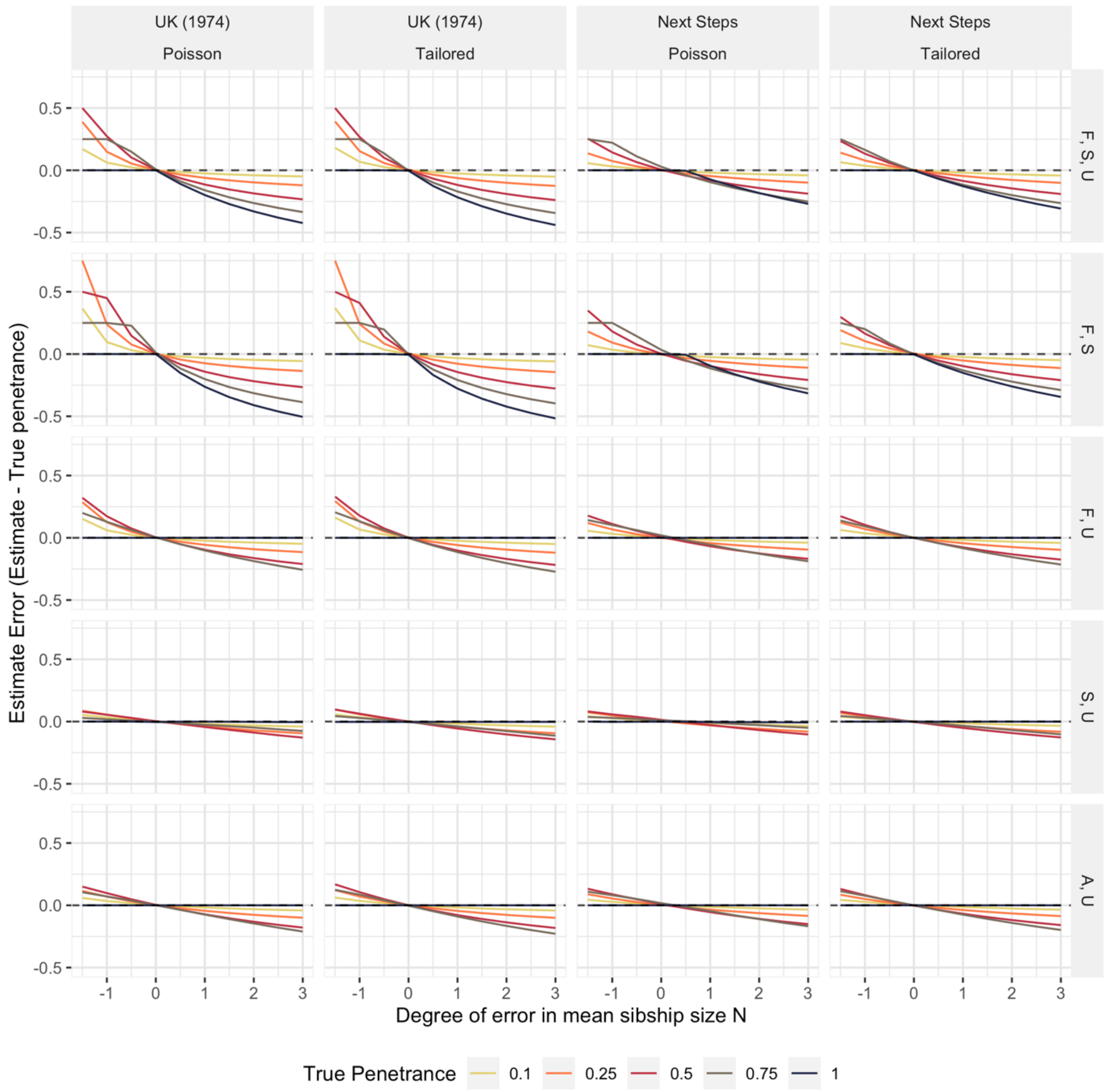


*Figure S5. Error in penetrance estimates according to degree of error in estimation of $N$. Zero indicates a perfect penetrance estimate, positive values indicate overestimation and negative values underestimation. Plot lines represent different true penetrance values. Panel columns stratify by population simulated (see Figure S3) and by Step 4 adjustment approach (see Supplementary Methods 1.1), which follows either the default approach (denoted Poisson) or is tailored to errors predicted under an internally-simulated sibship distribution directly approximating the sample data. Panel rows stratify estimates according to the disease state combination from which $R(X)^{obs}$ is defined; F = familial, S = sporadic, U = unaffected, and A = affected - state X is represented by the first state named.*

Misspecification of disease state rates*:*

This simulation study examines the accuracy of penetrance estimates when $R(X)^{obs}$ is incorrectly estimated. Several values of true penetrance were defined: $f_i^{true} = (0.10, 0.25, 0.50, 0.75, 1.00)$. A sequence of values to represent the degree of error in disease state rate estimates was also specified*:* $R(X)_i^{modify} = (-0.15, -0.10 \ldots, 15)$. The UK and NS populations were simulated as before. $R(X)^{obs}$ was calculated for a given $f_i^{true}$ across each of the five possible disease combinations, with the $R(X)^{obs}$ value to be defined in penetrance estimation being adjusted across each value of $R(X)_i^{modify}$; any adjusted $R(X)^{obs}$ values falling outside of the 0 to 1 interval were truncated to be 0.00001 or 1, according to whether they were below or that interval. Penetrance estimates, $f_i^{adjusted}$, were made for the simulated UK and NS populations, defining $N$ according to the mean sibship size of that sample, approximately 1.84 for the UK and 3.01 for the NS populations, and $R(X)^{obs}$ by the adjusted value obtained. For instance, if $R(X)^{obs} = 0.366$ and $R(X)_i^{modify} = 0.15$, then penetrance would be estimated based on $R(X)^{obs} = 0.516$. These results are presented in Figure S6.

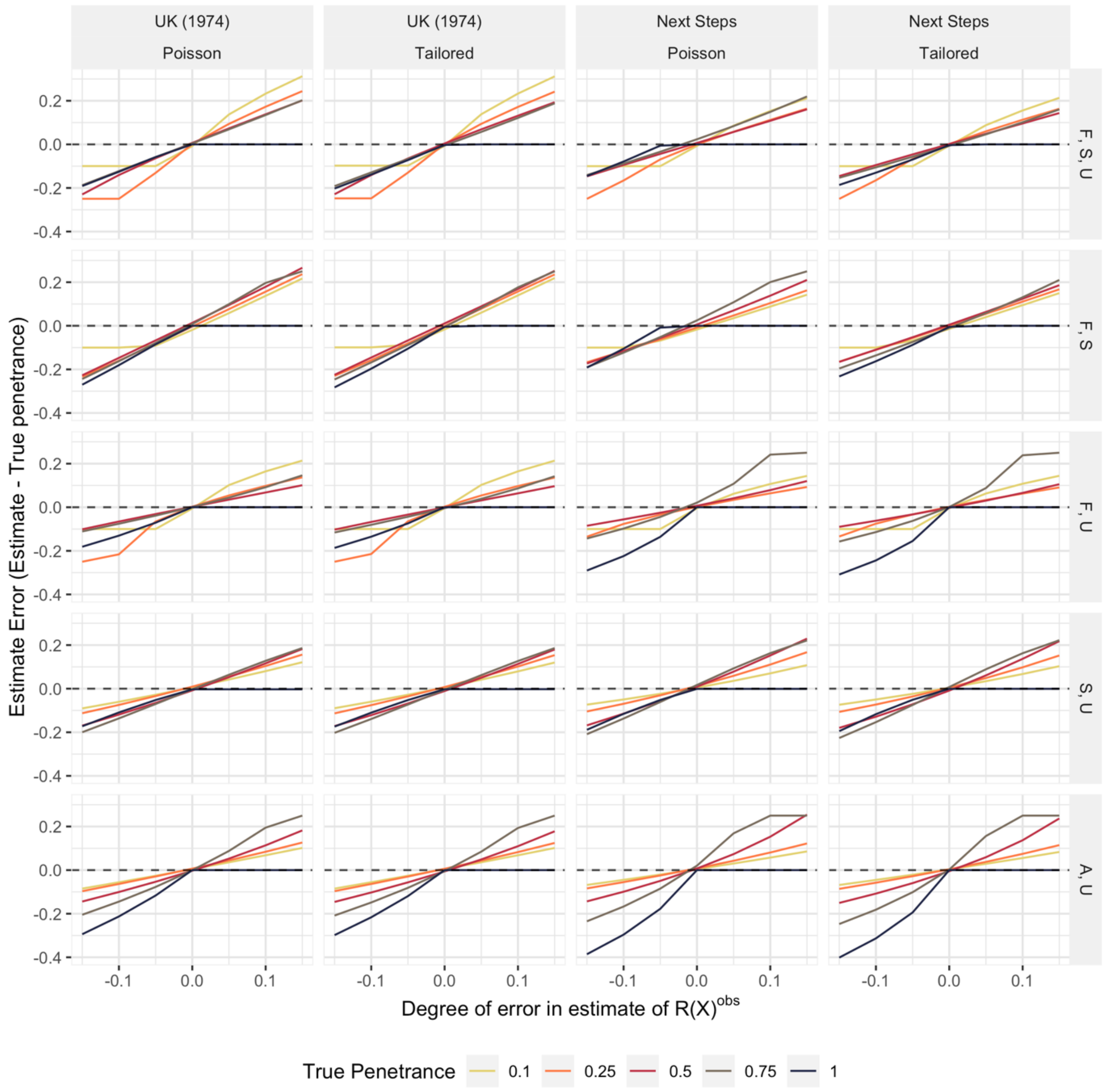


*Figure S6. Error in penetrance estimates according to degree of error in estimation of $R(X)^{obs}$. Zero indicates a perfect penetrance estimate, positive values indicate overestimation and negative values underestimation. Plot lines represent different true penetrance values. Panel columns stratify by population simulated (see Figure S3) and by Step 4 adjustment approach (see Supplementary Methods 1.1), which follows either the default approach (denoted Poisson) or is tailored to errors predicted under an internally-simulated sibship distribution directly approximating the sample data. Panel rows stratify estimates according to the disease state combination from which $R(X)^{obs}$ is defined; F = familial, S = sporadic, U = unaffected, and A = affected - state X is represented by the first state named.*

##### *1.3. ADPenetrance: a companion web tool*

This method of penetrance calculation is additionally available as an open-access web tool accessible at https://adpenetrance.rosalind.kcl.ac.uk. This was coded in R (Version 3.6.3) and leverages the R Shiny package (Version 1.4.0.2). An example of the interface and output of this tool is shown in Figure 2, as applied to estimation of *SOD1* variant penetrance for ALS using data from a European sample as described in case study 3.

This tool can be used calculate penetrance for a given variant based on an estimate of $R(X)^{obs}$ and a defined sibship size. State X is assigned to a particular state based on which disease states are included within input data, as indicated by the user. Those states represented can be any two or all three of the familial, sporadic, and unaffected states or the unaffected and affected states. If the familial state is represented within input data, then state $X$ is familial. If only the sporadic and unaffected states are represented, then state $X$ is sporadic. If the affected and unaffected states are represented, then state $X$ is affected.

The user can derive $R(X)^{obs}$ independently, manually specifying the rate of the state requested by the tool. Alternatively, they can provide variant characteristics and weighting factors (see Table 1), in order to calculate $R(X)^{obs}$ as described in Step 1. These variant characteristics can be given in each disease state as either (1) variant counts and sample size among population-based samples or (2) directly as variant frequencies.

If data are given using variant counts and sample sizes for each disease state, then the error propagation step is included by default, deriving the standard error for each variant frequency from these values. If data are given using variant frequencies or if $R(X)^{obs}$ is provided directly, then the user can opt to provide error terms for those estimates specified to enable error propagation. Error terms can be given either as standard errors or as confidence intervals from which standard errors are derived via z-score conversion. The user is asked to select which of these will be provided and, where confidence intervals are given, should indicate the level of confidence that these represent (95% confidence is assumed by default). Wherever error propagation is performed, the user will also need to specify the desired confidence level for the penetrance estimate output. This is to be selected from a series of options, where z-score conversion is used to transform the standard error of $R(X)^{obs}$ into the upper and lower confidence interval bounds of this estimate, which can then be used to estimate the bounds of the penetrance estimate.

The user must also indicate the average sibship size across the sample set. This can be specified either manually or by querying a repository of Total Fertility Rate estimates across many world regions which we have integrated within the tool[5].

Once input data are specified, the tool can be operated and $R(X)_i^{ex}$ is calculated for all values of $f_i$ between 0 and 1 at increasing increments of 0.0001. Penetrance is then estimated as in Steps 3 and 4 and a results table is produced.

The results Table presents $R(X)^{obs}$ and the estimated $R(X)_i^{ex}$, $f_i^{unadjusted}$, and $f_i^{adjusted}$ values to which this corresponds, additionally noting which state X represents. This $f^{adjusted}$ should be taken as the penetrance estimate. If error propagation is performed, upper and lower confidence intervals and the standard error of the $R(X)^{obs}$ will be provided, alongside corresponding confidence intervals for $R(X)^{ex}$,$f^{unadjusted}$, and $f^{adjusted}$.

#### 2. Supplementary Tables

##### *2.1. Table S1*

| Joint population[§] | | Population sampled | Number of unrelated people sampled | | | | Percentage of joint population[ø] | Calculation of sibship size ($N$) | | |
|---|---|---|---|---|---|---|---|---|---|---|
| | | | Sporadic | Familial | Unaffected | Total | | World region[*] | Weighted TFR estimate[†] | TFR Estimate for region[Δ] |
| European ancestry sample | North European | North American (white) | 2606 | 1450 | 4934 | 8990 | 50.20% | North America | 0.856381708 | 1.706 |
| | | British | 1145 | 192 | 1786 | 3123 | 17.44% | United Kingdom | 0.292961081 | 1.68 |
| | | German and Austrian | 803 | 231 | 436 | 1470 | 8.21% | Germany | 0.128868167 | 1.57 |
| | | Norwegian | 371 | 64 | 572 | 1007 | 5.62% | Norway | 0.08771679 | 1.56 |
| | | Australian | 578 | 252 | 0 | 830 | 4.63% | Australia | 0.080641018 | 1.74 |
| | | French | 300 | 174 | 348 | 822 | 4.59% | France | 0.086289575 | 1.88 |
| | | Swedish | 200 | 127 | 200 | 527 | 2.94% | Sweden | 0.05179072 | 1.76 |
| | | Irish | 236 | 35 | 212 | 483 | 2.70% | Ireland | 0.04719694 | 1.75 |
| | | Polish | 153 | 21 | 190 | 364 | 2.03% | Poland | 0.029674465 | 1.46 |
| | | Russian | 157 | 10 | 126 | 293 | 1.64% | Russian Federation | 0.025685968 | 1.57 |
| | | **Total** | **6549** | **2556** | **8804** | **17909** | 100% | - | - | **1.687206433**[Δ] |
| | South European | Italian and Sardinian | 2516 | 633 | 1040 | 4189 | 52.45% | Italy | 0.676660406 | 1.29 |
| | | Spanish | 806 | 283 | 544 | 1633 | 20.45% | Spain | 0.257648385 | 1.26 |
| | | Basque | 117 | 41 | 425 | 583 | 7.30% | Spain | 0.091983471 | 1.26 |
| | | Portuguese | 317 | 85 | 100 | 502 | 6.29% | Portugal | 0.089261207 | 1.42 |

| | | | | | | | | | | |
|---|---|---|---|---|---|---|---|---|---|---|
| | | Cretan | 174 | 92 | 0 | 266 | 3.33% | Greece | 0.044966191 | 1.35 |
| | | Serbian | 47 | 51 | 161 | 259 | 3.24% | Serbia | 0.048323316 | 1.49 |
| | | Greek | 235 | 0 | 0 | 235 | 2.94% | Greece | 0.03972577 | 1.35 |
| | | Chilean | 137 | 29 | 153 | 319 | 3.99% | Chile | 0.065869146 | 1.649 |
| | | **Total** | **4349** | **1214** | **2423** | **7986** | 100% | - | - | **1.314437891**[Δ] |
| | All European ancestry | North European | 6549 | 2556 | 8804 | 17909 | 69.16% | - | 1.166873142 | 1.687206433[Δ] |
| | | South European | 4349 | 1214 | 2423 | 7986 | 30.84% | - | 0.405371732 | 1.314437891[Δ] |
| | | **Total** | **10898** | **3770** | **11227** | **25895** | 100% | - | - | **1.572244874**[Δ] |
| Global ancestries sample | | Chinese | 1360 | 973 | 938 | 3271 | 9.55% | China | 0.161344638 | 1.69 |
| | | Japanese | 526 | 60 | 372 | 958 | 2.80% | Japan | 0.039704629 | 1.42 |
| | | Korean | 436 | 17 | 0 | 453 | 1.32% | Korean Republic | 0.012917547 | 0.977 |
| | | Indian | 718 | 82 | 1200 | 2000 | 5.84% | India | 0.12970638 | 2.222 |
| | | North African Arabs | 56 | 143 | 739 | 938 | 2.74% | Middle East and North Africa | 0.076902749 | 2.809 |
| | | Ashkenazi Jews | 259 | 78 | 410 | 747 | 2.18% | North America | 0.037195202 | 1.706 |
| | | All European ancestry | 10898 | 3770 | 11227 | 25895 | 75.58% | - | 1.188292598 | 1.572244874[Δ] |
| | | **Total** | **14,253** | **5,123** | **14,886** | **34,262** | 100% | - | **-** | **1.64606374**[Δ] |

*Table S1. Sample characteristics and calculation of N for data applied in case study 1[Ref. 6].*
*[§]Populations sampled were assigned to joint ancestry regions based on the ancestry group most frequent among people from that population*
*[*]Total Fertility Rate (TFR) estimates for each individual region drawn from the World Bank [5] database: these estimates were assigned to each population using the region defined in the World Bank database that appeared most representative of it. Where a sampled population features more than one named country, TFR was defined by the country with the larger population. For the Ashkenazi Jewish and Basque populations, estimates were assigned based on the world regions for which their populations are largest.*

[†]*Each weighted TFR value is calculated as:* $TFR\ estimate\ for\ region \times percentage\ of\ joint\ population;$
[Δ]*Marked TFR estimates were derived by summation of all w*eighted TFR estimates attributed to that region

#### *2.2. Table S2*

| | Sample | Variant frequency in familial state (variant count / sample size) | Variant frequency in sporadic state (variant count / sample size) | Variant frequency in unaffected state (variant count / sample size) | Average sibship size[†] | States modelled [#] | Familial disease rate among those harbouring the variant across states modelled (95% confidence interval) | Penetrance (95% Confidence interval) |
|---|---|---|---|---|---|---|---|---|
| Individual population estimates | North American (white) | 0.0310 (45/1450) | 0.00998 (26/2606) | $2.027\times10^{-4}$ (1/4934) | 1.706 | F, S | 0.267 (0.174, 0.361) | 0.436 (0.277, 0.596) |
| | Italian and Sardinian | 0.0411 (26/633) | 0.0147 (37/2516) | $9.615\times10^{-4}$ (1/1040) | 1.29 | F, S | 0.247 (0.155, 0.339) | 0.502 (0.311, 0.694) |
| | Spanish | 0.0495 (14/283) | 0.0273 (22/806) | 0 (0/544) | 1.26 | F, S | 0.175 (0.080, 0.270) | 0.361 (0.159, 0.562) |
| | Portuguese | 0.141 (12/85) | 0.0410 (13/317) | 0 (0/100) | 1.420 | F, S | 0.288 (0.135, 0.441) | 0.544 (0.246, 0.852) |
| | North African Arabs | 0.3566 (51/143) | 0.3929 (22/56) | 0.00541 (4/739) | 2.809 | F,S,U | 0.064 (0.036, 0.092) | 0.185 (0.135, 0.227) |
| | | | | | | F, S | 0.096 (0.062, 0.130) | 0.097 (0.060, 0.135) |
| | | | | | | F,U | 0.161 (0.026, 0.297) | 0.244 (0.101, 0.333) |
| | | | | | | S,U | 0.643 (0.407, 0.880)[ø] | 0.513 (0.254, 0.857) |
| | Ashkenazi Jews | 0.282 (22/78) | 0.0965 (25/259) | 0.00976 (4/410) | 1.706 | F,S,U | 0.063 (0.012, 0.115) | 0.254 (0.103, 0.355) |
| | | | | | | F, S | 0.255 (0.158, 0.353) | 0.416 (0.249, 0.582) |
| | | | | | | F,U | 0.078 (0.003, 0.152) | 0.232 (0.050, 0.317) |
| | | | | | | S,U | 0.197 (0.032, 0.363)[ø] | 0.127 (0.018, 0.263) |
| Joint population estimates | North European ancestry | 0.0274 (70/2556) | 0.00809 (53/6549) | $1.136\times10^{-4}$ (1/8804) | 1.687 | F, S | 0.284 (0.212, 0.356) | 0.469 (0.346, 0.592) |
| | South European ancestry | 0.0461 (56/1214) | 0.0177 (77/4349) | $4.127\times10^{-4}$ (1/2423) | 1.314 | F, S | 0.234 (0.173, 0.295) | 0.468 (0.343, 0.593) |
| | All European ancestry | 0.0334 (126/3770) | 0.0119 (130/10898) | $1.781\times10^{-4}$ (2/11227) | 1.572 | F, S, U | 0.170 (0.092, 0.249) | 0.468 (0.328, 0.587) |
| | | | | | | F, S | 0.247 (0.202, 0.292) | 0.429 (0.348, 0.509) |
| | | | | | | F, U | 0.354 (0.034, 0.673) | 0.494 (0.167, 0.709) |
| | | | | | | S, U | 0.625 (0.297, 0.952)[ø] | 0.547 (0.212, 0.950) |
| | Total Worldwide | 0.03923 (201/5123) | 0.01255 (179/14253) | $7.389\times10^{-4}$ (11/14886) | 1.646 | F, S, U | 0.098 (0.059, 0.137) | 0.332 (0.251, 0.402) |
| | | | | | | F, S | 0.268 (0.229, 0.307) | 0.450 (0.382, 0.517) |
| | | | | | | F, U | 0.134 (0.064, 0.204) | 0.304 (0.216, 0.370) |

| | | | | |
|---|---|---|---|---|
| | | S, U | 0.297 (0.170, 0.424)[ø] | 0.209 (0.110, 0.324) |

*Table S2. Penetrance estimation of the LRRK2 p.Gly2019Ser variant for Parkinson's Disease across populations sampled in case study 1. Population specific penetrance estimates were made only for those with at least 5 people harbouring LRRK2 p.Gly2019Ser in both the familial and sporadic states. Lifetime disease risk was 1/37 (0.027) in all calculations; the proportions familial and sporadic were respectively 0.105 and 0.895. The familial and sporadic states were modelled in all included populations. Penetrance was also modelled using the unaffected state in only the North African Arabic and Ashkenazi Jewish populations because the variant was sparse in all control samples – it was most frequent in these two groups.*

*[†]Estimated using Total Fertility Rates described for each population in Table S1[(Ref. 5)]; [#]F=familial, S=sporadic, U=unaffected (controls); [ø]Rate of sporadic disease has been calculated here because the familial state is not represented*

#### 2.3. *Table S3*

| *SOD1* variant | Variant frequency in familial state (variant count / sample size) | Variant frequency in sporadic state (variant count / sample size) | Variant frequency in unaffected state (variant count / sample size) | Lifetime risk of disease | Proportion familial* | Average sibship size† | States modelled# | Familial disease rate among those harbouring the variant across states modelled (95% confidence interval) | Penetrance (95% Confidence interval) ∞ |
|---|---|---|---|---|---|---|---|---|---|
| p.Ala5Val | 0.006222 (7/1125) | 0.000229 (1/4366) | - | - | 0.050 | 1.543 | F, S | 0.588 (0.081, 1§) | 1 (0.133, 1) |
| p.Asp91Ala¥ | - | 0.000916 (4/4366) | 0.00137 (33/24,143) | 0.0025 (Ref. 7) | 0.050 | 1.543 | S, U | 1.59x10$^{-3}$ (0.000, 0.003) ø | 8.98x10$^{-5}$ (0, 0.001) |
| p.Ile114Thr | 0.01491 (17/1140) | 0.001374 (6/4366) | - | - | 0.050 | 1.543 | F, S | 0.364 (0.149, 0.578) | 0.648 (0.256, 1) |

*Table S3. Penetrance estimation for widely described SOD1 variants. Variant frequencies are estimated using the ALS Variant Server[8] for the familial state, the ProjectMinE database[9] for the sporadic state and the European (non-Finnish) population of the gnomAD v2.1.1 (control) database[10] for the unaffected state.*

*Variant count = the number of people heterozygous for the tested variants; sample size = the number of people sequenced for variants at this locus; *Proportion sporadic is defined as 1 – proportion familial ($P(S|A) = 1 - P(F|A)$); †Estimated based on Total Fertility Rates for the European Union region in 2018(Ref. 5); #F=familial, S=sporadic, U=unaffected; §The familial disease rate estimate is truncated to 1 as the upper 95% confidence interval bound exceeds the highest possible frequency; ¥p.Asp91Ala is most frequently associated with autosomal recessive ALS presentations, we have modelled the penetrance of its autosomal dominant form only; øRate of sporadic disease has been calculated here because the familial state is not represented – no occurrences of the SOD1 p.Asp91Ala variant are reported in the ALS Variant Server. ∞Step 4 penetrance estimates are shown.*

#### 2.4. Table S4

| | | Phenotype | | Mathematical notation |
|---|---|---|---|---|
| | | ALS | FTD | |
| Published data | Lifetime risk (1/N) | 1/400 (Ref. 7) | 1/742 (Ref. 11) | A |
| | Familial disease rate (freq.) | 0.05 (Refs. 12, 13) | 0.30 (Ref. 13) | B |
| | *C9orf72*$^{RE}$ rate in familial state (freq.) | 0.32 (Ref. 14) | 0.248 (Ref. 15) | C |
| | *C9orf72*$^{RE}$ rate in sporadic state (freq.) | 0.05 (Ref. 14) | 0.060 (Ref. 15) | D |
| Estimated value$^{\Delta}$ | Overall *C9orf72*$^{RE}$ rate (freq.) | 0.064 | 0.116 | E$^{\#}$ |
| | Rate of *C9orf72*$^{RE}$ and phenotype in population (freq.) | $1.588 \times 10^{-4}$ | $1.568 \times 10^{-4}$ | F$^{\dagger}$ |
| | Incidence relative to FTD among people harbouring *C9orf72*$^{RE}$ | **1.012** | - | G$^{\emptyset}$ |

*Table S4. Estimation of the incidence of amyotrophic lateral sclerosis relative to frontotemporal dementia among people of European ancestry who harbour the pathogenic hexanucleotide GGGGCC repeat expansion of the C9orf72 gene (C9orf72$^{RE}$).*

$^{\Delta}$*Calculations are shown with respect to mathematical notation assigned to each row:*

$^{\#}E = (C \times B) + (D \times (1 - B))$;

$^{\dagger}F = E \times A$;

$^{\emptyset}G = F_{ALS}/F_{FTD}$.

#### 2.5. Table S5

| Case study | Data subset | States modelled # | Unadjusted penetrance estimates (95% Confidence interval) | | Adjusted penetrance (95% Confidence interval)∞ |
|---|---|---|---|---|---|
| | | | Lookup approach | Non-Linear Minimisation§ | |
| *LRRK2* p.G2019S for PD[6] | Total worldwide | F, S, U | 0.336 (0.258, 0.401) | 0.336 (0.258, 0.401) | 0.332 (0.251, 0.402) |
| | | F, S | 0.453 (0.394, 0.511) | 0.453 (0.394, 0.511) | 0.450 (0.382, 0.517) |
| | | F, U | 0.305 (0.220, 0.368) | 0.305 (0.220, 0.368) | 0.304 (0.216, 0.370) |
| | | S, U | 0.196 (0.103, 0.305) | 0.196 (0.103, 0.305) | 0.209 (0.110, 0.324) |
| *BMPR2* variants for PAH | All variants[16] | F, S | 0.395 (0.356, 0.433) | 0.395 (0.356, 0.433) | 0.382 (0.339, 0.426) |
| | All variants[17] | F, S | 0.302 (0.211, 0.390) | 0.303 (0.211, 0.390) | 0.281 (0.186, 0.376) |
| | Small variants[17] | F, S | 0.319 (0.204, 0.427) | 0.319 (0.204, 0.427) | 0.299 (0.179, 0.419) |
| | Large variants[17] | F, S | 0.243 (0.023, 0.439) | 0.243 (0.023, 0.439) | 0.218 (0.014, 0.432) |
| *SOD1* variants for ALS[18] | Asian | F, S | 0.749 (0.629, 0.864) | 0.749 (0.629, 0.864) | 0.829 (0.665, 1) |
| | European | F, S | 0.660 (0.494, 0.812) | 0.660 (0.494, 0.812) | 0.705 (0.496, 0.933) |
| *C9orf72*$^{RE}$ for ALS[14] | Asian | F, S | 0.282 (0.023, 0.514) | 0.282 (0.023, 0.514) | 0.263 (0.016, 0.522) |
| | European | F, S | 0.449 (0.377, 0.518) | 0.449 (0.377, 0.518) | 0.443 (0.363, 0.524) |

*Table S5. Comparison of unadjusted penetrance estimates derived for the case studies presented in Table 2 between the lookup table and maximum-likelihood approaches. #F=familial, S=sporadic, U=unaffected (controls); C9orf72$^{RE}$ = the pathogenic C9orf72 GGGGCC hexanucleotide repeat expansion; §This approach is described in Supplementary Methods 1.2.1; ∞These adjusted penetrance estimates were derived using the unadjusted penetrance estimate obtained using the lookup approach.*
